# Chronic pain: transdiagnostic meta-analytic evidence of convergent network signature with PTSD

**DOI:** 10.64898/2026.01.21.26344503

**Authors:** Menghan Li, Yaxian Hou, Liu Dan, Yali Zhou, Mercy Chepngetich Bore, Jianuo Lei, Junjie Wang, Michelle Hei Lam Tsang, Michael Maes, Keith M. Kendrick, Benjamin Becker, Stefania Ferraro

**Author notes:** Corresponding author: Stefania Ferraro.

## Abstract

Chronic pain is increasingly conceptualized within a stress-related framework. However, it remains unclear whether chronic pain and prototypical stress-related conditions—such as post-traumatic stress disorder (PTSD)—share common neurobiological substrates. To this end, we conducted a pre-registered transdiagnostic meta-analytic study of gray matter volume alterations in chronic pain (60 studies) and PTSD (20 studies). Disorder-specific meta-analyses revealed that chronic pain was associated with distributed volume reductions across ventromedial prefrontal, middle cingulate, and insular cortices, whereas PTSD exhibited a single cluster of reduced volume in the anterior cingulate/dorsomedial prefrontal cortices. A conjunction analysis revealed that both conditions converged onto an overlapping cluster of reduced volume in the bilateral medial orbitofrontal/anterior cingulate area. Using normative resting-state fMRI data (HCP 7T dataset), we found that chronic pain neuroanatomical abnormalities were embedded within a distributed architecture of large-scale circuits encompassing mesocorticolimbic/reward, default mode, salience, frontoparietal, dorsal attention, and somatosensory networks. On the other hand, the PTSD focal neuroanatomical alteration was embedded in a single large-scale circuit mapping onto the mesocorticolimbic/reward, default mode, salience, and visual networks. In both conditions, the mesocorticolimbic/reward circuit emerged as the most robustly involved large-scale network. Notably, the shared cluster of reduced volume showed functional integration within the mesocorticolimbic/reward and default mode networks, with neurochemical fingerprinting revealing robust spatial correspondence with dopaminergic, serotonergic, opioid, and endocannabinoid receptor/transporter maps. Overall, these findings suggest that chronic pain and PTSD, beyond disorder-specific alterations, converge on a shared large-scale network organization. The overlap between chronic pain and a prototypical stress-related disorder at the network level provides neurobiological support for conceptualizing chronic pain within a stress-related framework.

## 1. Introduction

Chronic pain (ChP) represents one of the most prevalent and disabling health conditions worldwide, affecting up to one-third of the adult population (Jackson et al., 2016; Rometsch et al., 2025; Sá et al., 2019). The condition is defined by the International Association for the Study of Pain (IASP) as “pain that persists or recurs for more than three months” (Treede et al., 2019). While this temporal criterion is essential for clinical and operational purposes, it is important to recognize that pain exists on a dynamic continuum, ranging from an adaptive neurobiological response to a chronic and maladaptive state, in which neuroplastic changes encompass molecular and synaptic modifications to large-scale network reorganization. Over the past decade, our understanding of ChP has undergone substantial progress. Once considered primarily as an amplification of nociceptive activity driven by peripheral and central inflammation and central sensitization (Woolf, 2011; Latremoliere and Woolf, 2009; Basbaum et al., 2009; Ji et al., 2014), ChP is now increasingly conceptualized within a stress-related framework(Vachon-Presseau, 2018), with a major impact on mesocorticolimbic circuits (Thierry et al., 1976; Cabib and Puglisi-Allegra, 1996; Quessy et al., 2021; Baik, 2020; Martucci et al., 2018; Ferraro et al., 2022, 2025), where it influences a wide range of cognitive and affective processes, including reward, aversion, motivation, and emotions.

At the neurobiological level, stress represents an orchestrated and multifaceted set of physiological and neurochemical responses to internal or external challenges —whether real or anticipated—that are perceived as threatening or harmful to the individual (Buenrostro-Jáuregui et al., 2025). These responses are primarily regulated by the paraventricular nucleus (PVN) of the hypothalamus in concert with the mesocorticolimbic system (Vachon-Presseau, 2018). The PVN engages multiple interacting pathways with reciprocal feedback that span hypothalamic–pituitary–adrenal (HPA) axis activity (Timmers et al., 2019), autonomic regulation (Yeater et al., 2021), neuroactive steroid signaling, and immune processes (Woda et al., 2016; McEwen et al., 2016). The mesocorticolimbic circuits encompass the ventral tegmental area (VTA), the ventral striatum (comprising the nucleus accumbens), as well as the amygdala, the hippocampus, and prefrontal regions including the medial prefrontal cortex. Within these circuits the hippocampus and the prefrontal cortices directly modulate stress responses through their glutamatergic projections to the PVN, within a feedback loop that operates in concert with both glutamatergic and GABAergic inputs from the amygdala (De Kloet et al., 2005). Prolonged stress exerts its effects on the HPA axis in two successive stages (Agorastos and Chrousos, 2022). In the early phase, individuals often display hyperactivation of the HPA axis, characterized by increased corticotropin-releasing hormone (CRH) and adrenocorticotropic hormone (ACTH) release, hypercorti-solemia, and impaired glucocorticoid negative feedback—a profile sometimes referred to as ‘cortisol resistance’ (Miller et al., 2007). This condition has been associated with reduced glucocorticoid receptor expression in limbic regions, particularly the medial prefrontal cortex and hippocampus (Gómez et al., 1996; Mizoguchi et al., 2003). Over time, this hyperactive profile can transition to a downregulated state, characterized by lower baseline cortisol, heightened feedback sensitivity to glucocorticoids (Juruena et al., 2020; Heim et al., 2000; Raison and Miller, 2003), and marked plastic changes in mesocorticolimbic forebrain structures. The hippocampus, which is rich in glucocorticoid and mineralocorticoid receptors (McEwan et al., 1968; Reul and Kloet, 1985), is particularly vulnerable to chronic stress, showing dendritic atrophy and impaired neurogenesis (McEwen et al., 2016). Pronounced neuroplastic changes in dendritic architecture are also observed within the basolateral and medial amygdala, resulting in hyperactivity that disrupts the balance between contextual regulation and emotional reactivity (McEwen et al., 2016; Snyder et al., 2011; Sapolsky, 1996; Vyas et al., 2002; Roozendaal et al., 2009). Beyond these structures, chronic stress also disrupts the mesocorticolimbic pathways mainly involved in motivation and reward processing. Neuroplastic alterations were observed within the VTA–basolateral amygdala–nucleus accumbens circuit with reduced dopaminergic signaling (Holly and Miczek, 2016; Baik, 2020). These alterations extend to the prefrontal cortex (Lee and Goto, 2011), further weakening top–down regulation and exacerbating emotional dysregulation, thereby fostering anhedonia and increasing vulnerability to psychiatric disorders (Arnsten, 2009; McEwen and Morrison, 2013). Collectively, these findings converge within the allostatic model (McEwen, 1998; Koob and Le Moal, 2001; Koob and Schulkin, 2019), which conceptualizes chronic stress as a driver of reward dysregulation, affective disturbances, and addiction vulnerability.

The relatively recent reconceptualization of ChP as a disorder of mesocorticolimbic circuits (Baliki et al., 2010, 2012; Vachon-Presseau et al., 2016a) reveals a striking convergence with what is known about the effects of chronic stress on these same systems. Consistently, ChP is characterized by robust anatomical and functional alterations of these circuits: structural abnormalities in the hippocampus, amygdala, and nucleus accumbens; reduced dopaminergic signaling in the VTA and nucleus accumbens with blunting of reward responsiveness; and impaired prefrontal control (Baliki et al., 2012; Taylor et al., 2016; Borsook et al., 2016; Serafini et al., 2020; Finan and Smith, 2013; Martikainen et al., 2015; Vachon-Presseau et al., 2016b; Yang and Chang, 2019; Ferraro et al., 2022, 2025). The link between ChP and stress is further supported by the evidence that this disorder is characterized by multidimensional alterations in stress-regulatory systems—including dysregulation of the HPA axis, autonomic imbalance, immune–neuroendocrine dysfunctions (Woda et al., 2016), and increased basal levels of cortisol (Vachon-Presseau et al., 2013; Borsook et al., 2012), manifesting clinically as stress intolerance (Wyns et al., 2023).

Yet, the precise role of chronic stress in ChP disorders remains incompletely understood, with the underlying shared neural pathways and their interactions remaining largely unexplored. Disentangling these mechanisms is central to our understanding of ChP, not only for conceptual clarity but also for the development of novel therapeutic approaches. A significant step forward in this direction is to test whether ChP carries a stress-related neural signature. To this end, a promising strategy is a transdiagnostic meta-analytic approach contrasting studies investigating ChP conditions with studies investigating PTSD, a prototypical stress-related disorder. PTSD is a psychiatric condition typically developed after a single or repeated traumatic event (e.g., violence, warfare, abuse), yet its symptoms may persist for months or even years, effectively transforming the traumatic experience into a state of prolonged internal distress. According to the Diagnostic and Statistical Manual of Mental Disorders, Fifth Edition (DSM-5) classification, PTSD is now considered a trauma- and stressor-related disorder and is no longer classified as an anxiety disorder (Friedman et al., 2011). Moreover, from a neurobiological perspective, PTSD can be conceptualized as a prototypical state of chronic stress, characterized by HPA axis dysregulation, amygdala hyperactivity, reduced hippocampal and amygdala volume (Logue et al., 2018), and prefrontal cortical dysfunction. Supporting the rationale for a transdiagnostic comparison between ChP and PTSD, epidemiological studies have shown that a substantial proportion of ChP individuals also meet criteria for PTSD (10–50%) (Asmundson et al., 2002; Carty et al., 2011; Morgan and Aldington, 2020; Noel et al., 2016), and vice versa, with estimates suggesting that 20–80% of PTSD individuals also suffer from ChP (Asmundson et al., 2002; Otis et al., 2009). This reciprocal comorbidity indicates a shared vulnerability and mutual maintenance, and may even reflect manifestations of a unitary underlying process, in which the mesolimbic system with learning mechanisms might play a central role (Abdallah and Geha, 2017). Such a meta-analytic transdiagnostic approach might enable the identification of overlapping stress-related signatures within ChP morphological alterations, while also highlighting specific ChP alterations. Crucially, conducting this comparison within a meta-analytic framework is essential to integrate heterogeneous findings across studies and to isolate, with the highest level of available evidence, the impact of chronic stress on pain-related brain alterations. By leveraging case-control voxel-based morphometry (VBM) studies in patients with ChP and PTSD, this pre-registered metaanalysis directly compares morphometric alterations in ChP and PTSD patients to delineate shared neural signatures that may reflect stress-related mechanisms in ChP, as well as distinct signatures that might instead capture disorder-specific abnormalities. Furthermore, these findings were embedded into large-scale functional networks and complemented them with cognitive decoding and neurotransmitter mapping, thereby moving from focal structural abnormalities to a systems-level perspective. Accordingly, we implemented a multi-step transdiagnostic meta-analytic framework. Specifically, we first identified robust gray matter volume abnormalities in the chronic pain and PTSD datasets using separate coordinate-based meta-analyses, and then defined the overlap of these abnormalities using a conjunction analysis. Second, we examined condition-specific brain alterations across the ChP and PTSD datasets using meta-regression analyses. Third, we conducted a pooled meta-analysis including all ChP and PTSD studies to quantify the degree of heterogeneity associated with each cluster of abnormalities. Finally, we contextualized the resulting clusters in terms of large-scale functional network organization, cognitive decoding, and neurotransmitter receptor and transporter distributions.

We hypothesized that ChP and PTSD patients would share a common set of abnormal brain regions, primarily localized within mesocorticolimbic systems, with particular involvement of limbic structures and their prefrontal regulatory regions. In addition, we hypothesized that ChP and PTSD would differ in the involvement of other brain networks, particularly those supporting sensory processing.

## 2. Materials and Methods

### 2.1. Studies selection

This meta-analysis was pre-registered on the Open Science Framework (https://osf.io/m2bzu/overview) and was conducted in accordance with the PRISMA 2020 reporting guidelines (see Figures 1 and 2). To identify relevant studies, an extensive search was conducted in the following databases: PubMed, Web of Science, and Scopus. The following separate search queries were used for each condition to retrieve studies published between 2000 and 2024: 1) ChP: (“VBM” OR “voxel-based morphometry” OR “morphometry” OR “thickness” OR “grey matter”) AND (“chronic pain” OR “neuropathic” OR “fibromyalgia” OR “irritable bowel syndrome”); 2) PTSD: (“VBM” OR “voxel-based morphometry” OR “morphometry” OR “thickness” OR “grey matter”) AND (“PTSD” OR “posttraumatic stress disorder”). The search results for each condition were merged across databases, and duplicates were removed. Each eligible study was given a unique code. Two reviewers (S.F. for ChP and M.L. for PTSD) excluded records not relevant for the current meta-analysis based on the titles and abstracts. A third reviewer (Y.H.) verified the consistency of the selection. A record was excluded at this stage only if both the initial reviewer (S.F. or M.L.) and the third reviewer agreed on its exclusion. The selected studies were then assessed by M.L. via fulltext review to identify records meeting the inclusion criteria and then independently cross-checked by S.F. and Y.H. Studies were included if they fulfilled the following criteria: (1) published in a peer-reviewed journal in English; grey literature and non-English publications were excluded, (2) investigated brain morphometric differences between patients (ChP or PTSD) and a healthy control group, (3) used voxel based morphometry (VBM) with a whole-brain approach, (4) investigated only adult participants between 18 and 65 years old, (5) investigated at least 10 participants per group, (6) reported specific peak foci of brain morphometric differences in either Talairach and Tournoux (TAL) (Talairach, 1988) or Montreal Neurological Institute (MNI) space (Evans et al., 1993), (7) did not report results already presented in other included studies, (8) did not report differences between groups. For the selection of the ChP records, we included studies investigating patients that had a diagnosis of a pain condition considered chronic regardless of a time criterion, and we excluded studies investigating pain conditions associated with cortical reorganization unrelated to chronic pain per see, such as spinal cord injury, tumors of any origins, chronic pancreatitis (Muthulingam et al., 2019), and medication overuse headache (Sun et al., 2025). Furthermore, we excluded dysmenorrhea and episodic migraine, since, although recurrent pain conditions, they cannot be considered chronic. For the PTSD dataset, we included studies investigating patients who had a formal diagnosis. Importantly, large-scale consortium analyses such as ENIGMA-PTSD (Wang et al., 2021) were not included in the quantitative meta-analysis because they aggregate voxel-wise results from multiple cohorts, many of which have already been published as independent studies. Including these results would therefore violate study-level independence, while their harmonized outputs do not provide the coordinate-level information required for inclusion in coordinate-based meta-analytic models. For each selected article, M.L. extracted the statistical threshold and the stereotactic space (Talairach or MNI) used, the peak coordinates of the significant clusters identified in the group comparison, as well as the corresponding z- or t-values. When statistical values were not reported, the corresponding entries were marked as “NA”. Other additional information, including the number of participants with mean age, sex distribution, and the software employed for the analyses, was also collected. To exclude the presence of potential extraction errors or other issues, such as the inclusion of studies based on regions of interest (ROIs), S.F. independently rechecked the selected studies and the extracted coordinates from both databases. In case of discrepancies, the study or the coordinates were re-examined collegially and corrected as needed. Notably, for the ChP dataset, when studies reported multiple ChP conditions (e.g., chronic back pain and complex regional pain syndrome) compared to a single CTRL group, we reported the aggregated results (i.e., all ChP conditions vs. CTRL) when a combined analysis was provided. If only separate results were available for each condition, we included them individually in the meta-analysis and treated them as dependent samples. For the PTSD dataset, when studies included comparisons between PTSD patients and both healthy controls and trauma-exposed healthy controls, we selected the results from the comparison between PTSD patients and trauma-exposed healthy controls.

**Figure 1.**
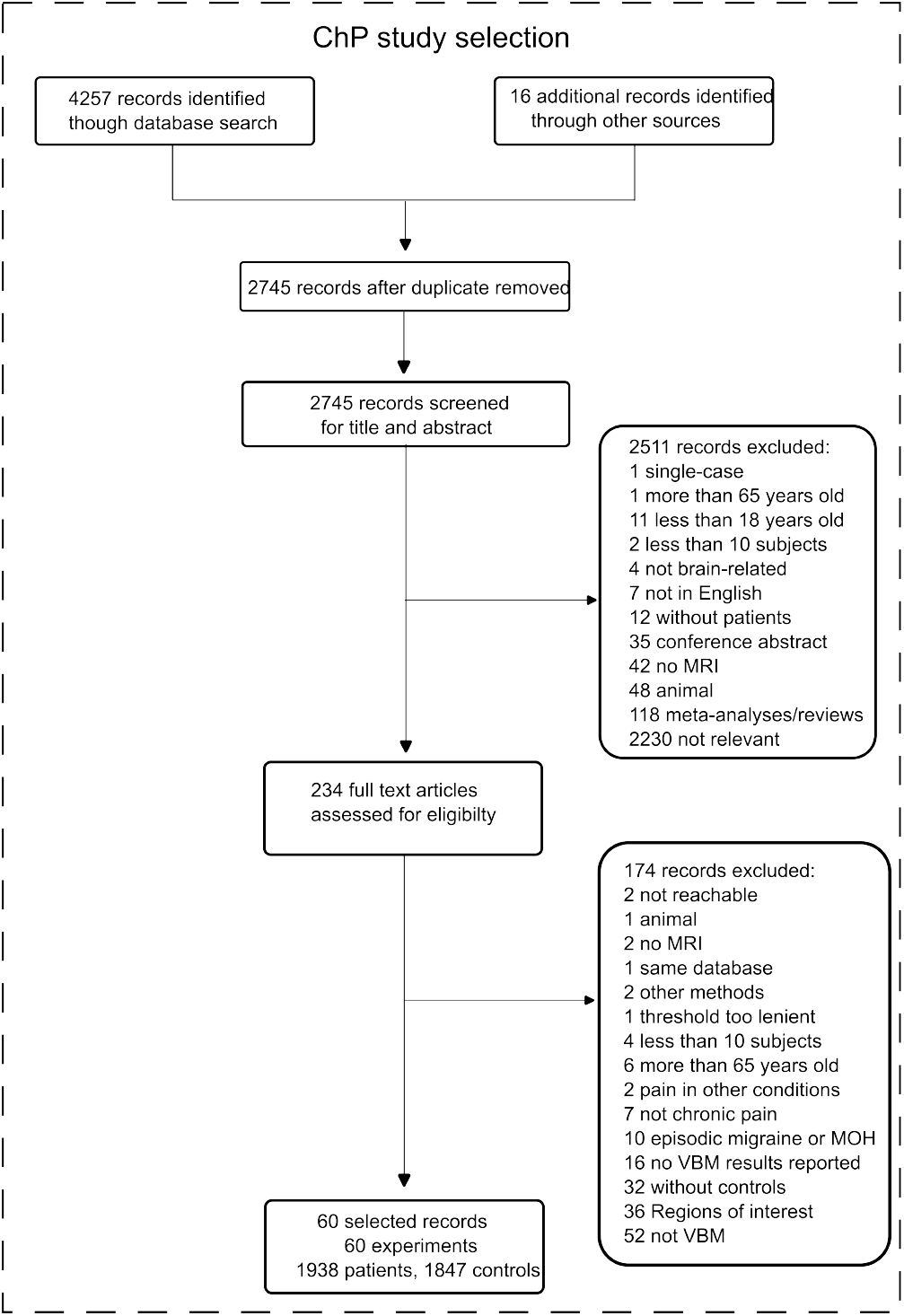
Flowchart of the screening process of the chronic pain (ChP) studies dataset according to the PRISMA guideline.

**Figure 2.**
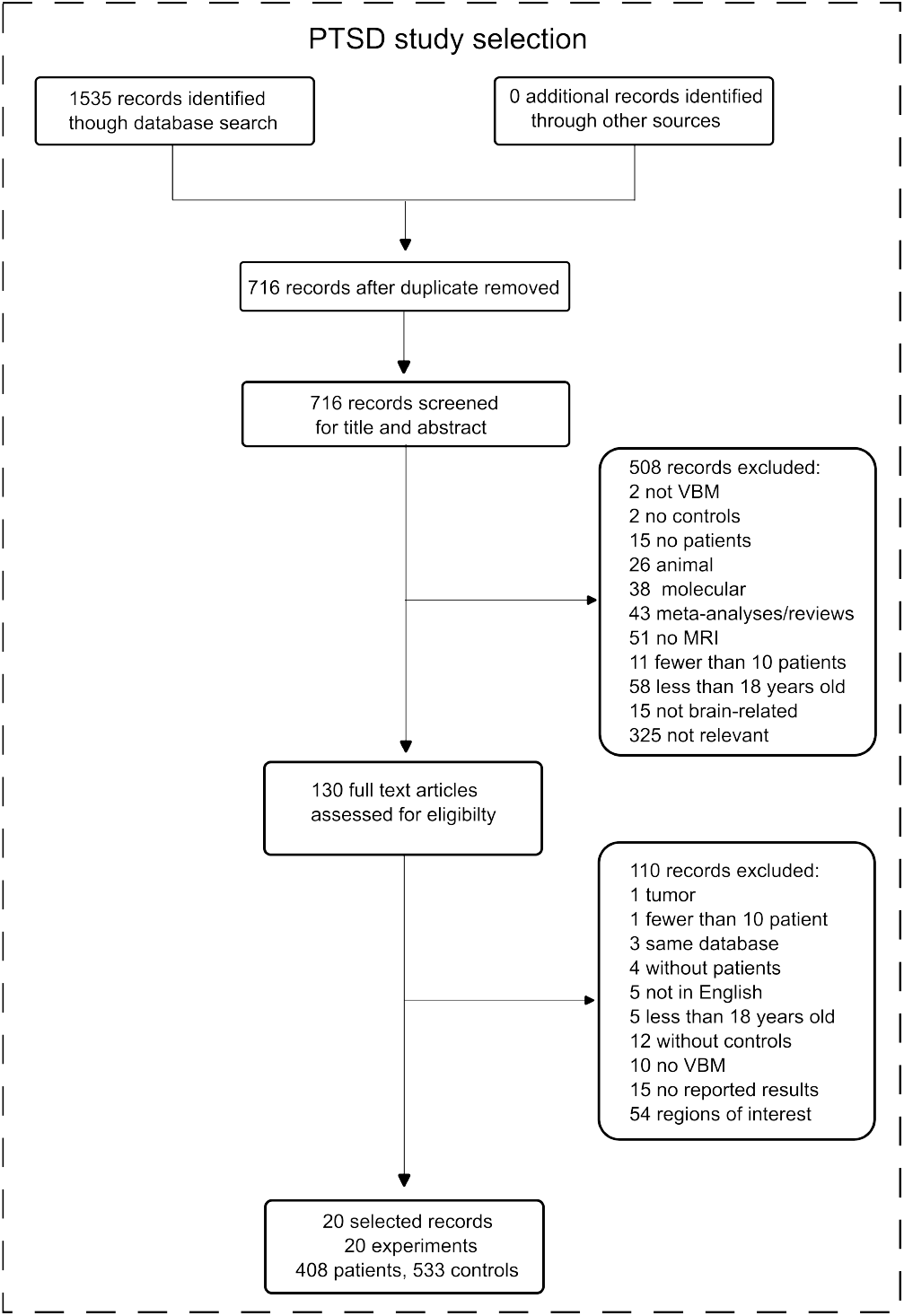
Flowchart of the screening process of the post-traumatic stress disorder (PTSD) studies dataset according to the PRISMA guideline.

### 2.2. Meta-analyses

All meta-analyses were conducted using Seed-based d Mapping with Permutation of Subject Images (SDM-PSI, 6.23v - https://www.sdmproject.com), after converting z-values to t-values where appropriate. The obtained maps were thresholded at *p* < 0.05 TFCE (threshold-free cluster enhancement)-corrected for multiple comparisons, with a minimum cluster size of *k* > 10 voxels. For all the conducted meta-analyses and for each significant cluster, we calculated the effect size (measured using Hedges’ g), the between-study heterogeneity, estimated using *τ* ^2^ and expressed as the *I*^2^ index (*I*^2^ > 50% indicating substantial heterogeneity) (Martins et al., 2021), and Egger’s test to determine the potential publication bias (*p* < 0.05). To identify specific and common alterations between the two conditions, we adopted the following approach.

First, we delineated consistent patterns of regional abnormalities associated with ChP or with PTSD, performing two separate meta-analyses, one for each condition (hereafter, ‘ChP meta-analysis’ and ‘PTSD metaanalysis’). Furthermore, we identified convergent volumetric abnormalities between the two disorders by performing a conjunction (Conj) analysis —using the minimum statistic approach (Jakobs et al., 2012; Nichols et al., 2005)—on the thresholded maps resulting from the ChP and PTSD meta-analyses. This yielded a Conj map generated using SPM12.

Second, we identified condition-specific (i.e., divergent) brain alterations between the ChP and PTSD datasets (hereafter, ‘condition-specific meta-regression’) by performing a meta-regression (linear model), as implemented in SDM-PSI. This model included all studies from both datasets, and group membership was entered as a dummy-coded variable (1 = ChP, 0 = PTSD).

Third, since heterogeneity represented a key focus of our comparative framework, we performed a pooled metaanalysis including all the studies from the ChP and PTSD datasets (hereafter, ‘pooled meta-analysis’). This allowed us to directly quantify the overall degree of heterogeneity across the two conditions by calculating the *I*^2^ index for each significant cluster.

### 2.3. Seed-based functional connectivity of the identified altered clusters

Using an unbiased approach, we characterized the spatial patterns of functional connectivity of the abnormal brain regions identified in the ChP and PTSD metaanalyses, as well as the convergent regions of alteration identified by the Conj analysis. To this aim, we examined their seed-based functional connectivity (SBC) employing the resting-state functional magnetic resonance imaging (rsfMRI) data from 30 subjects (age: M = 29.2 ys, SD = 3.32 ys; 20 females) of the publicly available Human Connectome Project dataset [HCP - Young Adult; for details https://www.humanconnectome.org/hcp-protocols-ya-7t-imaging (Smith et al., 2013; Van Essen et al., 2013)] using the 4 rsfMRI runs acquired at 7T (900 volumes per run; 1.6 mm isotropic voxels; TR = 1000 ms; TE = 22.2 ms; flip angle = 45°; FOV = 208 × 208 mm; Smith et al., 2013; Ugurbil et al., 2013), and already preprocessed and available as ‘rfMRI *hp2000 clean.nii.gz’. These images were fed into the CONN v21a toolbox (), where they were smoothed using a 4 mm full-width at half-maximum (FWHM) Gaussian kernel and denoised following a standard preprocessing pipeline (Nieto-Castanon, 2020). To conduct the SBC analyses, the seeds were defined as 6-mm radius spherical regions centered on the peak coordinates of the identified clusters. Whole-brain functional connectivity strength was computed as Fisher-transformed bivariate correlation coefficients, derived from a weighted general linear model estimated separately for each seed region and target voxel. In the Group-level analyses, a separate GLM was estimated for each voxel, with first-level connectivity measures at this voxel as dependent variables. Voxel-level hypotheses were evaluated using multivariate parametric statistics with random-effects across subjects and sample covariance estimation across multiple measurements. Inferences were performed at the level of individual clusters (groups of contiguous voxels). Cluster-level inferences were based on parametric statistics from Gaussian Random Field theory. Results were thresholded using a cluster-forming voxel level threshold (*p* < 0.001) combined with a cluster-extent threshold corrected for multiple comparisons (*p* < 0.05, FDR-corrected), yielding the seed-based connectivity (SBC) maps (hereafter, ‘SBC maps’).

### 2.4. Functional characterization of the seed-based functional connectivity maps

To determine to which canonical rs-fMRI network each SBC map belonged, we used a null map-based analytical framework (Burt et al., 2020). As canonical rsfMRI networks, we employed the functional networks available in the CONN toolbox: namely, the salience, sensorimotor, visual, dorsal-attention, fronto-parietal, and the default mode (DMN) networks. Moreover, given our strong a priori hypothesis of mesocorticolimbic system involvement (Holly and Miczek, 2016; Baik, 2020, Lee and Goto, 2011), we employed Neurosynth to produce a robust map of this circuit. Neurosynth is an open platform that automatically generates term-based meta-analyses from a large database of neuroimaging studies (at the time of the current analyses, including 14,371 studies and 1,334 terms). In line with previous literature, we used the term ‘reward’ in Neurosynth to capture the mesocorticolimbic system, given that reward processing is consistently regarded as the core function mediated by this circuit ((Haber and Knutson, 2010; Berridge and Kringelbach, 2015). Building on this correspondence, a term-based meta-analysis of the term ‘reward’ (identifying 922 studies) was performed, generating a map (hereafter referred to as mesocorticolimbic/reward network) through an association test based on z-scores derived from a two-way ANOVA assessing the relationship between term usage and voxel activity. The association test determines whether activation in a voxel occurs more consistently in studies mentioning the term of interest compared to those that do not, thereby generating a co-activation map specifically linked to the term of interest. To assess the significance of overlap between the SBC maps and each canonical network, we generated 1000 null maps for each canonical network (Burt et al., 2020). Because spatial autocorrelation, typical of brain MRI data, violates the assumptions of value permutation and parametric testing (Markello and Misic, 2021), null maps were generated using the algorithm in Burt (Burt et al., 2020), as implemented in Neuromaps (Markello et al., 2022). We then computed the overlap between the SBC map of interest and these null maps to create a null distribution for each canonical network. A p-value was obtained by calculating the proportion of overlap scores greater than the observed overlap, testing whether the overlap was greater than expected by chance. We considered overlaps significant at *p* < 0.001 (empirical permutation test; 1,000 permutations)).

### 2.5. NiMARE decoding of the seed-based functional connectivity maps

To explore the cognitive functions and anatomical structures associated with each produced SBC map, we used NiMARE’s decoding tool (https://nimare.readthedocs.io/en/stable/decoding.html). This tool estimates the cognitive relevance of brain maps by computing spatial correlations between the input map and a large database of meta-analytic co-activation maps. For each map, the decoder provides a ranked list of cognitive terms along with their corresponding correlation values. For interpretability, we arbitrarily selected the top 40 terms. If the same term was represented in different forms (e.g., “orbitofrontal” and “OFC”), we retained only the first occurrence. Moreover, two authors (S.F. and M.L.) independently reviewed these terms and identified those that referred to cognitive processes or brain regions, excluding the others. Any disagreements were resolved by consensus. As a post-hoc analysis, to assess whether the identified terms were shared across conditions, we aligned the full list of terms across all SBC maps (with each row corresponding to the same term) and computed second-order correlations (i.e., correlations between the r values obtained from the decoding procedure) between the ChP and PTSD SBC maps.

### 2.6. Neurotransmitter receptors/transporters profiling of the seed-based functional connectivity maps

As the final step, we investigated the neurotransmitter receptor profiling of the SBC maps. To achieve this, we used the JuSpace toolbox (version 2.0;) (Dukart et al., 2021), and computed the spatial correlations between these functional maps and the normative receptor/-transporter distribution templates derived from PET studies. These PET maps, linearly rescaled for comparability, are based on previously published datasets collected from healthy control samples. Using an a priori approach, we selected the dopaminergic, serotonergic, noradrenergic, opioid, and endocannabinoid systems as the target maps given their key involvement in the ChP and PTSD conditions (Ney et al., 2021; Huang et al., 2019; Finn et al., 2021; Nakamoto and Tokuyama, 2023; Krystal and Neumeister, 2009; Millan, 2002; Lopez-Martinez et al., 2019; Neumeister et al., 2013; Aby et al., 2022; Ravenelle et al., 2025). As multiple maps are available in JuSpace for each neurotransmitter system, we reduced the risk of type I error choosing the most representative ones for each system. Thus, we selected the FDOPA map (Gómez et al., 2018) as a representative marker of the dopaminergic system; the SERT map (Savli et al., 2012; Fazio et al., 2016; Beliveau et al., 2017) to capture the serotonergic system; the mu-opioid receptor (MOR/MU) maps (Kantonen et al., 2020; Turtonen et al., 2021) and the kappa-opioid receptor (KOR/KappaOp) maps as indices of the opioid system; the CB1 map (Normandin et al., 2015; Laurikainen et al., 2019) to represent the endocannabinoid system; and the NAT map (Hesse et al., 2017) for the noradrenergic system. All results were corrected for multiple comparisons (*p* < 0.05, Benjamini–Hochberg FDR-corrected).

## 3. Results

### 3.1. ChP and PTSD Datasets characteristics

Concerning the ChP dataset (see Table 1, and in SM, Tables S1 and S2), we identified 60 studies (totalling 65 experiments) reporting structural brain differences between 1938 patients (age, M *±* = 48 *±* 8.3, 1261 females among studies reporting sex distribution; patients in each study: median 23, range: 10-305) and 1847 CTRL subjects (age, M *±* = 46 *±* 8.3, 1064 females for studies with available sex distribution; number of subjects in each study: median 22, range: 10-296). Among these records, the majority employed corrected thresholds at the voxel or cluster level, or used an uncorrected voxel-wise threshold of *p* < 0.001. However, 8 studies (Ceko et al., 2013; Niddam et al., 2017; Pomares et al., 2017; Schweinhardt et al., 2008; Tang et al., 2021; Tsai et al., 2018; Wood et al., 2009; Kuchinad et al., 2007) applied more lenient thresholds, such as voxel-wise forming thresholds of *p* < 0.01 or *p* < 0.05 before cluster correction, or AlphaSim correction *p* < 0.05.

**Table 1:** Descriptive statistics and group comparisons of demographic variables for the ChP and PTSD datasets. *The computation of the percentage of female patients in each dataset is based on the studies that reported sex distribution. Abbreviations: ChP, chronic pain; PTSD, post-traumatic stress disorder.

|  | Participants | Age (M ± SD) | Females / Total subjects* |
| --- | --- | --- | --- |
| # Patients |  |  |  |
| ChP dataset | 1938 | 48 ± 8.3 | 1261 / 1930 (65.3%) |
| PTSD dataset | 408 | 41 ± 12.5 | 179 / 355 (50.4%) |
| Statistic | - | t(82)= 4.56; $p < 0.001$ | $\chi^2(1)=28.62;p < .0001$ |
| # Controls |  |  |  |
| ChP dataset | 1847 | 46 ± 8.3 | 1064 / 1807 (58.9%) |
| PTSD dataset | 533 | 46 ± 13.7 | 210/ 469 (44.8%) |
| Statistics | - | t(74) = 3.82; $p < 0.001$ | $\chi^2(1) = 31.90; p < 0.001$ |

Among the selected records, 3 studies (Baliki et al., 2011; Sundermann et al., 2019; Tsai et al., 2018) compared different ChP disorders with the same CTRL group, and the results were therefore considered dependent. In contrast, one study (Ceko et al., 2013) investigated two different age groups of patients with a ChP disorder, each matched with distinct control groups. The results were therefore considered independent. Being aware that any classification is a simplification of complex clinical realities, we classified the ChP conditions examined in the selected studies according to the mechanistic pain descriptors adopted by the International Association for the Study of Pain (IASP): nociceptive, nociplastic, and neuropathic (Raja et al., 2020). In particular, acknowledging that most ChP conditions may involve multiple pathophysiological mechanisms, we assigned each condition to the descriptor considered most representative of its predominant mechanism. With this in mind, the ChP dataset comprised 17 studies investigating neuropathic pain, 29 focused on nociplastic pain, and 16 on nociceptive pain. Additionally, 3 studies examined multiple types of ChP disorder. Concerning the PTSD dataset (see Tables 1, and in SM, Tables S3 and S4), we identified 20 studies reporting structural brain differences between 408 patients (age, M *±* SD =41 *±* 12.5, 179 females among studies reporting sex distribution; patients in each study: median 17, range: 10-57) compared to 533 CTRL subjects (age, M *±* SD = 46 *±* 13.7, 210 females; number of subjects in each study: median 19, range: 10-163). All the selected studies employed corrected thresholds at the voxel or cluster level, or used an uncorrected voxel-wise threshold of *p* < 0.001. Notably, the ChP and PTSD datasets differed significantly in both age and sex distribution (see Table 1). The ChP studies included older patients and a higher proportion of women (65.3% vs. 50.4%), compared to the PTSD studies. Please note that not all studies reported sex distribution (missing in 3 ChP and 4 PTSD records).

### 3.2. Meta-analyses

See Figure 3 and Table 2 for the results of this section. The ChP meta-analysis revealed that patients, compared to CTRL subjects, presented 4 clusters of reduced grey matter volume. A broad bilateral cluster was observed within the bilateral ventromedial prefrontal regions encompassing the frontal medial orbital gyrus and the gyrus rectus (peak at [–6, 48, –6]). Additional clusters were found in the bilateral middle cingulate cortex (peak at [2, −30, 40]), in the anterior section of the right insula (peak at [32, 16, –12]) extending to the right orbital gyrus, and in the posterior section of the left insula comprising the left Rolandic operculum (peak at [–50, –20, 12]). No evidence of increased regional grey matter in ChP patients compared to CTRL subjects was observed.

**Table 2:**
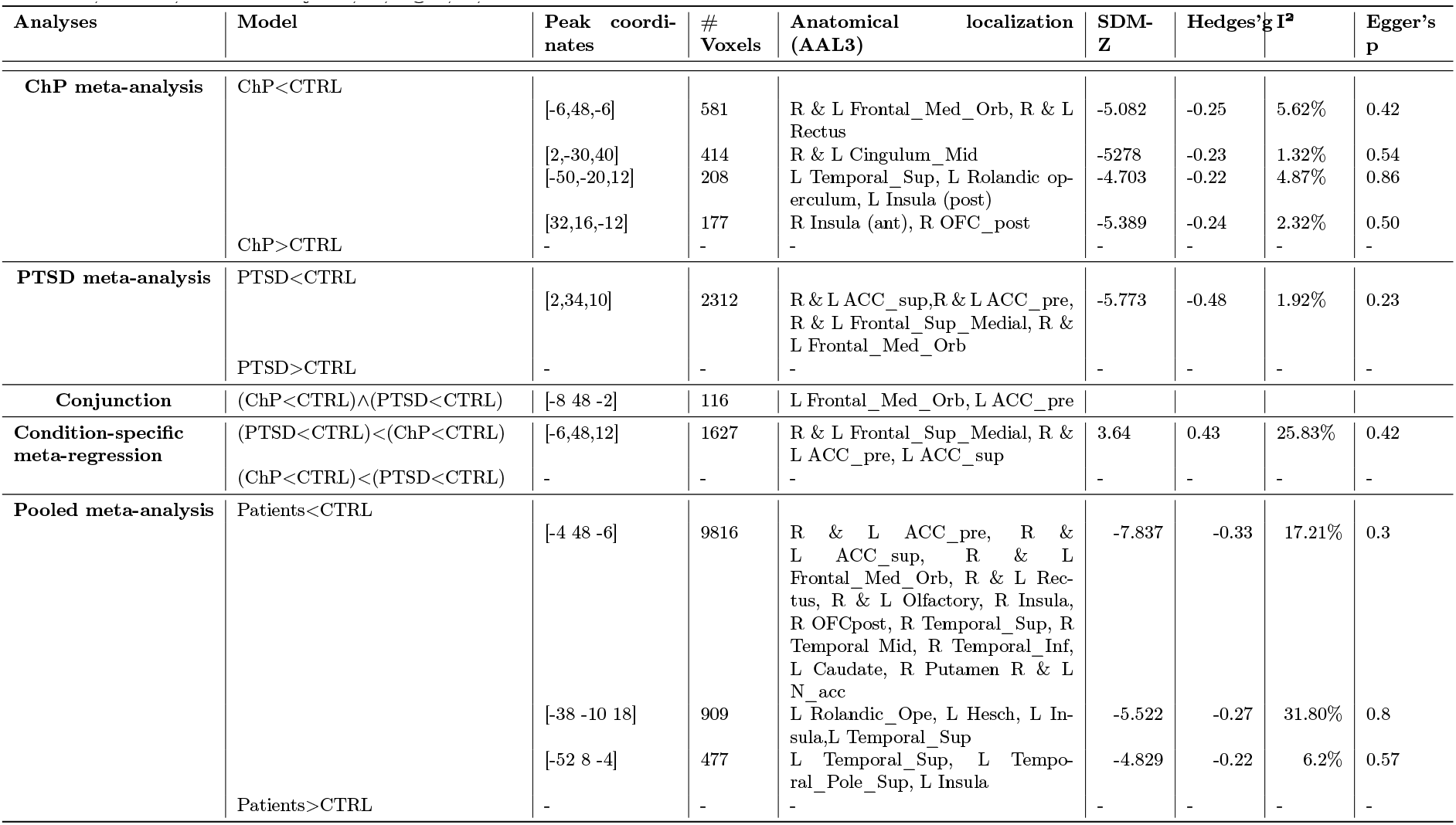
Results and anatomical localization of clusters of abnormalities identified by the meta-analyses and the conjunction analysis. Regions are reported using original AAL3 (Rolls et al., 2020) labels to avoid potential misnomers or inconsistencies. All reported coordinates are expressed in the MNI standard space. Abbreviations: ChP, chronic pain; PTSD, post-traumatic stress disorder; CTRL, control subjects; R, right; L, left.

**Figure 3.**
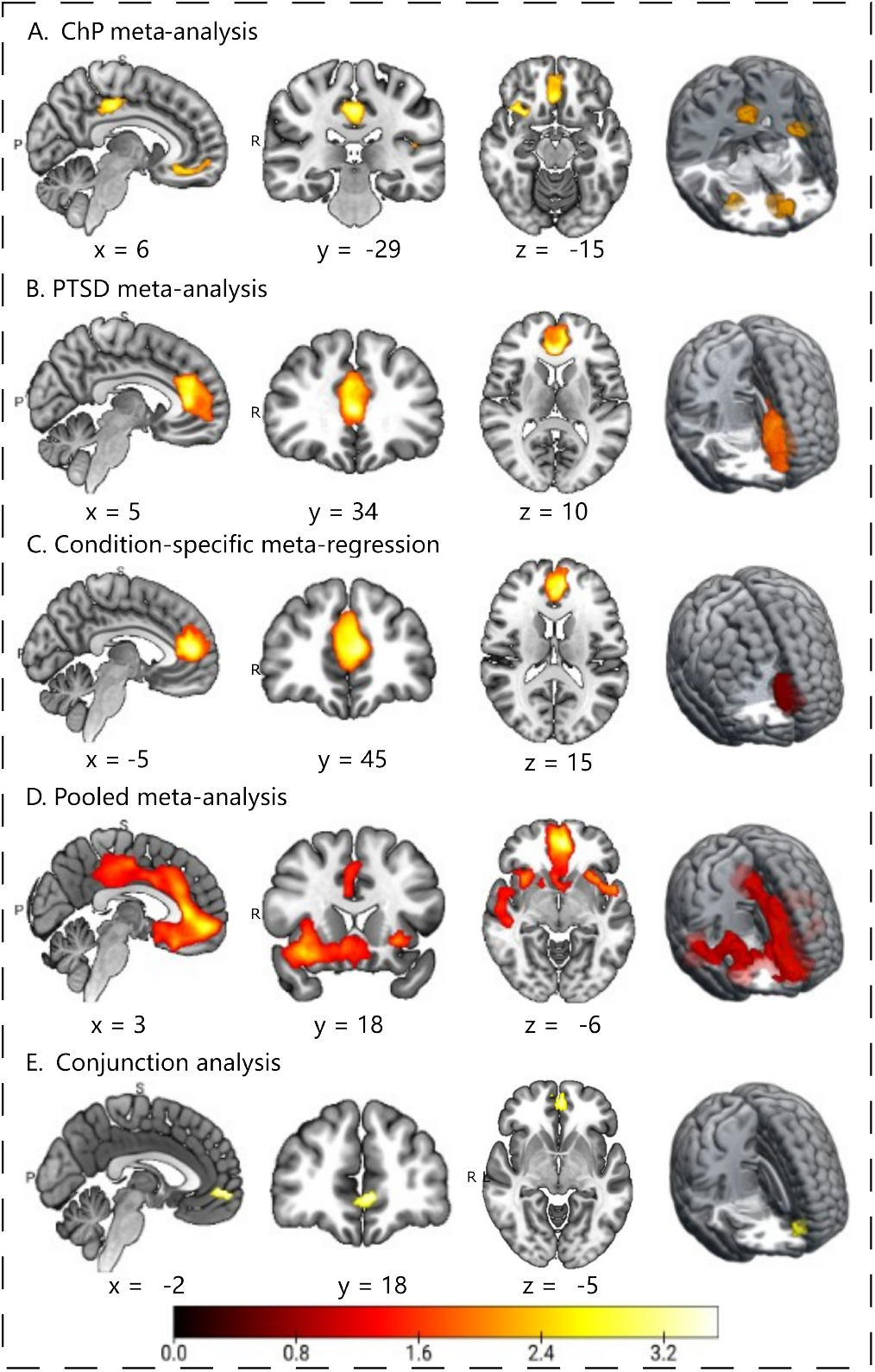
Results of SDM-based meta-analyses and conjunction analyses conducted on voxel-based morphometry (VBM) studies investigating differences between controls and patients with ChP or PTSD. For all the presented meta-analyses results, voxel-wise SDM-Z maps were thresholded at p < 0.05 TFCE-corrected for multiple comparisons, with a minimum cluster size of k > 10 voxels. A. Maps showing significant gray matter volume differences derived from the ChP dataset (60 VBM studies), reflecting brain regions where controls exhibit greater gray matter volume than ChP patients (controls > patients). B. Maps showing significant gray matter volume differences derived from the PTSD dataset (20 VBM studies), reflecting brain regions where healthy controls exhibit greater gray matter volume than PTSD patients (controls > patients). C. Maps showing significant gray matter volume differences derived from a condition-specific meta-regression comparing ChP and PTSD datasets, highlighting regions where ChP patients exhibit greater gray matter volume than PTSD patients (ChP > PTSD). D. Pooled meta-analysis maps including ChP and PTSD studies, showing brain regions where controls exhibit greater gray matter volume than patients (controls > patients). E. Maps showing brain regions identified by the conjunction analysis of the ChP and PTSD meta-analytic results, highlighting areas of overlapping gray matter volume reduction shared by both conditions. Coordinates are reported in Montreal Neurological Institute (MNI) space. Abbreviations: ChP, chronic pain; PTSD, post-traumatic stress disorder.

The PTSD meta-analysis revealed that patients presented a single extended bilateral cluster of decreased grey matter volume covering the anterior cingulate cortex, comprising the supracallosal and pregenual anterior cingulate cortex (peak at [2, 34, 10]), and the dorsomedial prefrontal regions, encompassing the frontal medial orbital and superior frontal gyri. The heterogeneity measures for the significant clusters observed in both meta-analyses were relatively low, with *I*^2^ values ranging between 2% and 6%. The Conj analyses of CP and PTSD meta-analytic maps revealed a single common cluster of decreased grey matter volume (116 voxels) in the bilateral frontal medial orbital gyrus (peak in [−8 48 −2]; centroid in [−2, 50, −6]).

The meta-regression assessing divergent brain alterations between the two datasets (i.e., condition-specific metaregression) showed that PTSD patients, in respect to ChP patients, presented decreased regional volumes localized in one cluster covering the bilateral superior frontal gyrus (medial section, comprising the dorsomedial prefrontal cortex) and the anterior cingulate cortex (peak: [−6, 48, 12]). Contrary to our expectations, no decrease in regional volumes was observed in the ChP dataset with respect to the PTSD dataset.

The pooled meta-analysis, including data from both ChP and PTSD studies, revealed 3 large clusters of decreased gray matter volume comprising the bilateral medial prefrontal regions, the cingulate cortex, the anterior and posterior areas (comprising the Rolandic operculum) of the insula cortex, and several subcortical areas, including the bilateral nucleus accumbens, a key component of the mesolimbic circuit. In this case, as expected, the *I*^2^ values increased, but never reached thresholds indicative of substantial heterogeneity (*I*^2^ ranging between 6% and 32%). Notably, the cluster identified at MNI coordinates [−52 8 −4]-encompassing the left insula (both anterior and posterior portions) and the Rolandic operculum—exhibited low levels of heterogeneity, with an *I*^2^ value of only 6.2%. Remarkably, in all the meta-analyses performed—whether on individual datasets, condition-specific or pooled meta-analyses — the Egger’s tests revealed no evidence of publication bias.

### 3.3. Functional characterization of the clusters of interest: seed-based functional connectivity maps

See Figure 4, and SM (Tables S5, S6, S7, S8, S9, S10) for the results of this section. Concerning the results of the ChP meta-analysis, the SBC maps seeded in 3 out of the 4 clusters of altered volumetry (peaks in frontal medial orbital cortex; [–6, 48, –6]), middle cingulate cortex; [2, −30, 40], and right anterior insula cortex [32, 16, –12]) revealed robust functional connectivity, with bilateral medial prefrontal areas, the precuneus, hippocampus, and nucleus accumbens. Conversely, the fourth cluster—peak in the left posterior insula [−50 −20 12])-exhibited strong connectivity with bilateral sensorimotor cortices, left posterior insular cortex, and bilateral thalamus. Concerning the PTSD meta-analysis, the SBC map of the observed cluster of altered volumetry (centered in the anterior cingulate cortex, peak at [2 34 10]) demonstrated robust bilateral functional connectivity with medial prefrontal areas, anterior and posterior insular cortices, middle cingulate cortex, and precuneus. Subcortical connectivity was also strong, involving the thalamus, hippocampus, hypothalamus, and nucleus accumbens. The SBC map seeded in the region of converging grey matter volume reduction observed in the Conj analysis (i.e., the overlap between ChP and PTSD datasets) and located in the bilateral frontal medial orbital gyrus revealed that this area was a node of an extended network primarily involving the bilateral medial regions of the prefrontal cortex—including the anterior cingulate cortex, the superior frontal medial gyrus, and the frontal medial orbital gyrus—as well as several subcortical structures such as the bilateral nucleus accumbens, hippocampus, parahippocampal gyrus, and the hypothalamus. Additionally, it showed strong connectivity with the precuneus, the medial thalamic nuclei, and cerebellar areas.

**Figure 4.**
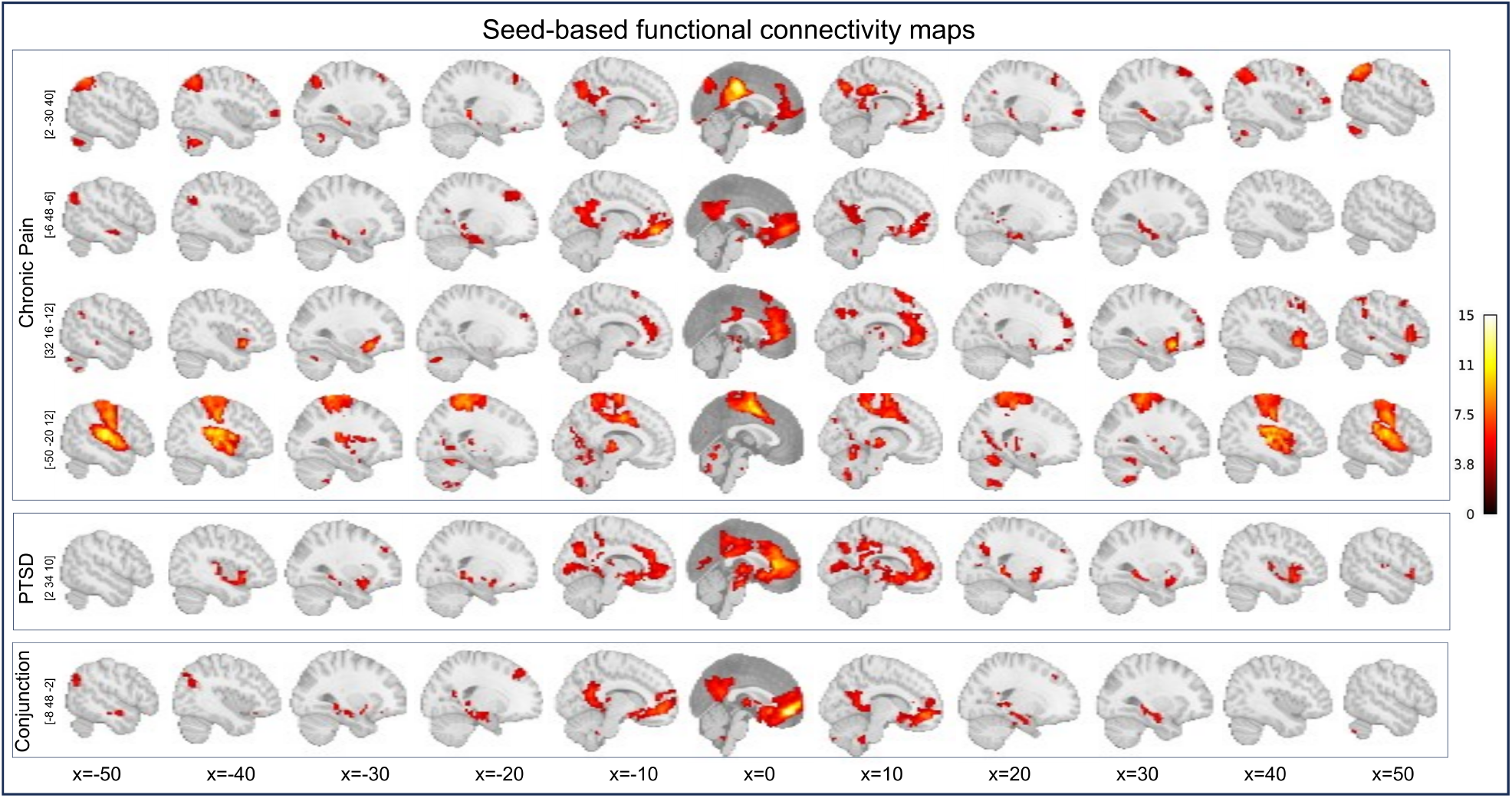
Graphical representations of SBC maps, shown as T-statistic maps, seeded in the peak coordinates obtained from the ChP and PTSD meta-analyses and the conjunction analysis. These maps were generated using resting-state functional magnetic resonance imaging (rs-fMRI) data from a subset of 30 participants (age: M = 29.2 years, SD = 3.32; 20 females) drawn from the publicly available Human Connectome Project dataset (HCP–Young Adult; for details see https://www.humanconnectome.org/hcp-protocols-ya-7t-imaging). Color bar represents T-statistics. Abbreviations: Conj, conjunction analysis; ChP, chronic pain; PTSD, post-traumatic stress disorder; SBC, seed-based connectivity analysis.

### 3.4. Seed-based resting-state functional connectivity maps: identification of the canonical rs-fMRI network of belonging

See Table 3 and Figure 5 for the results of this section. In the ChP dataset, the different SBC maps constituted large-scale functional circuits that mapped onto different canonical networks with distinct co-occurrence patterns. The SBC map seeded in the middle cingulate cortex at [2, −30, 40] showed significant overlap with the DMN, frontoparietal, and mesocorticolimbic/reward networks, whereas the SBC map seeded in the frontal medial orbital cortex at [−6, 48, −6] overlapped with the DMN and mesocorticolimbic/reward network. The SBC map seeded in the right anterior insula at [32, 16, −12] showed overlap with the frontoparietal, salience, and mesocorticolimbic/reward networks. By contrast, the SBC map seeded in the left posterior insula at [−50, −20, 12] was associated with both dorsal attention and sensorimotor networks.

**Table 3:** Formal tests assessing the similarity between SBC maps—centered on clusters of reduced regional grey matter volume from the ChP and PTSD meta-analyses and the conjunction analysis—and canonical rs-fMRI networks. All reported coordinates are expressed in the MNI standard space.* Significant results (*p* < 0.000). Abbreviations: DMN, default mode network; DAN, dorsal-attention network, SAL, salience network; ChP, chronic pain; PTSD, post-traumatic stress disorder; rs-fMRI, resting-state functional magnetic resonance imaging; SBC, seed-based connectivity; emp, empirical overlap value derived from null maps.

| Canonical fMRI networks |  |  |  |  |  |  |  |  |  |  |  |  |  |  |  |
| --- | --- | --- | --- | --- | --- | --- | --- | --- | --- | --- | --- | --- | --- | --- | --- |
| rs-fMRI networks |  |  |  |  |  |  |  |  |  |  |  |  |  | Co-activation network |  |
|  |  | DMN |  | DAN |  | FrontoParietal |  | SAL |  | Sensorimotor |  | Visual |  | Reward |  |
| | SBC map at | $p$ | $emp$ | $p$ | $emp$ | $p$ | $emp$ | $p$ | $emp$ | $p$ | $emp$ | $p$ | $emp$ | $p$ | $emp$ |
| ChP dataset | [2 -30 40] | 0.000* | 0.671 | 1.000 | 0.357 | 0.000* | 0.859 | 0.994 | 0.465 | 0.871 | 0.501 | 0.442 | 0.516 | 0.000* | 0.585 |
|  | [-6 48 -6] | 0.000* | 0.704 | 0.980 | 0.487 | 1.000 | 0.428 | 0.966 | 0.484 | 0.023 | 0.541 | 0.080 | 0.530 | 0.000* | 0.635 |
|  | [32 16 -12] | 0.105 | 0.542 | 1.000 | 0.434 | 0.000* | 0.622 | 0.000* | 0.615 | 1.000 | 0.435 | 0.144 | 0.535 | 0.000* | 0.636 |
|  | [-50 -20 12] | 1.000 | 0.461 | 0.000* | 0.718 | 0.661 | 0.565 | 0.017 | 0.639 | 0.000* | 1.048 | 0.995 | 0.544 | 0.997 | 0.535 |
| PTSD dataset | [2 34 10] | 0.000* | 0.629 | 1.000 | 0.444 | 1.000 | 0.450 | 0.000* | 0.701 | 0.950 | 0.509 | 0.000* | 0.577 | 0.000* | 0.743 |
| Conj analysis | [-8 48 -2] | 0.000* | 0.701 | 0.995 | 0.469 | 1.000 | 0.354 | 1.000 | 0.438 | 0.076 | 0.528 | 0.034 | 0.530 | 0.000* | 0.647 |

**Figure 5.**
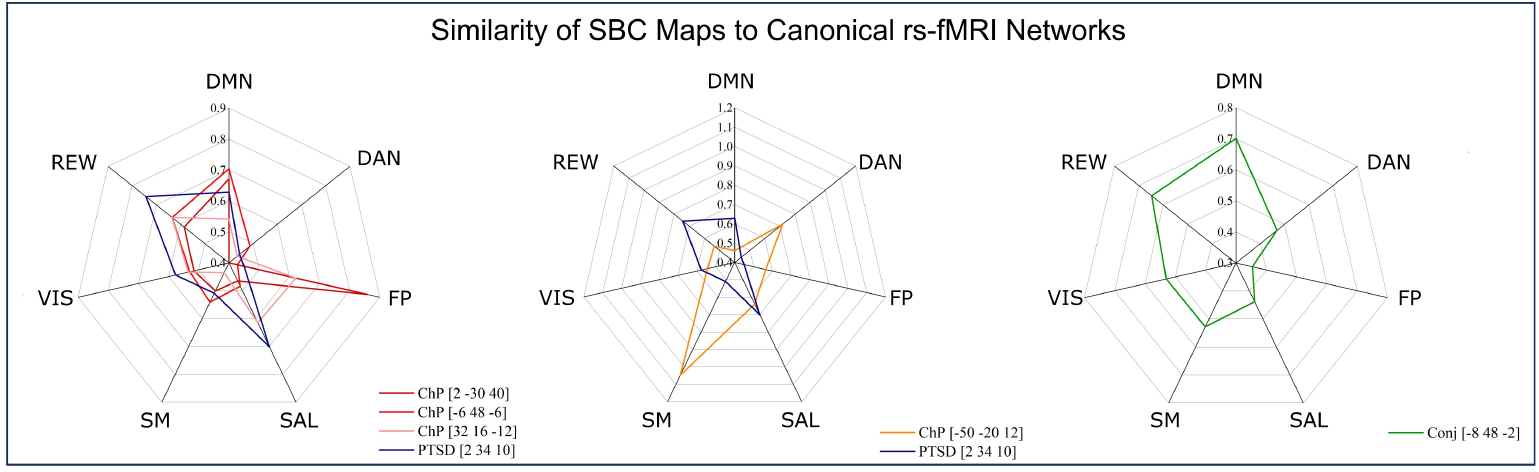
Radar charts showing the similarity between SBC maps —seeded at the peak coordinate (in brackets) of reduced regional grey matter volume from the ChP and PTSD meta-analyses and the conjunction analysis- and canonical rs-fMRI networks. For clarity, the SBC maps are displayed separately: (A) Radar chart for the ChP SBC maps and the PTSD SBC map, excluding the ChP map mapping onto the sensorimotor cortex; (B) Radar chart of the ChP and PTSD SBC maps mapping onto the sensory cortices; (C) Radar chart of the Conj analysis SBC map. Black dots indicate regions of significant overlap as determined by the null map approach. Abbreviations: SBC, seed-based functional connectivity; rs-fMRI, resting-state fMRI; ChP, chronic pain; PTSD, post-traumatic stress disorder; Conj, conjunction; SBC, seed-based functional connectivity; DMN, default mode network; DAN, dorsal attentional network; FP, frontoparietal network; SAL, salience network; SM, sensorimotor network; VIS, visual network; Reward, mesocorticolimbic/reward network.

Overall, in the ChP dataset, SBC maps were predominantly associated with the mesocorticolimbic/reward network (3 out of 4 SBC maps), which co-occurred with the DMN either alone or in combination with the frontoparietal network, as well as with the frontoparietal network in combination with the salience network. In contrast, the SBC map centered on the left posterior insula, associated with both the dorsal attention and sensorimotor networks, appeared to constitute a distinct system, separable from the reward-mediated network configuration observed in the other SBC maps. In the PTSD dataset, the single disorder-specific SBC map seeded in the anterior cingulate cortex at [2 34 10] exhibited simultaneous overlap with multiple large-scale systems, specifically the DMN, mesocorticolimbic/reward, salience, and visual networks. Notably, the mesocorticolimbic/reward network showed the highest empirical overlap (empirical overlap = 0.743), followed by the salience network (empirical overlap = 0.701). Taken together, these findings indicate that the mesocorticolimbic/reward network is the most consistently involved large-scale system across both datasets, being most recurrent in ChP and showing the strongest empirical overlap in PTSD. Moreover, both conditions mapped onto the mesocorticolimbic/reward, DMN, and salience networks, whereas frontoparietal, dorsal attention, and sensorimotor networks were specific to ChP and the visual network to PTSD. Consistent with this pattern, the SBC map derived from the conjunction analysis (seeded in the frontal medial orbital gyrus at [–8 48 –2]) showed robust overlap with the DMN and mesocorticolimbic/reward network.

### 3.5. Seed-based resting-state functional connectivity maps: NiMARE decoding

See Table 4 and Figure 6 for the results of this section. The decoding of the SBC maps derived from 3 out of the 4 clusters identified in the ChP meta-analysis ([2 −30 40], [−6 48 −6], and [32 16 −12]), as well as from the single cluster identified in the PTSD meta-analysis and from the Conj analysis (i.e., the region of convergent gray matter volume reductions across the two datasets), revealed highly similar terms. Indeed, despite the different cluster locations, the associated terms consistently referred to reward related processes (“reward”), affective processing (“face” and/or “affective”), autonomic regulation (“autonomic”), and memory functions (“memory”, “semantic”), together with anatomical terms related to the limbic system (“amygdala”, “hippocampus”, “OFC”). Importantly, we observed very high correlations between the PTSD term profile and the term profile of the SBC maps derived from the 2 ChP clusters seeded at [2 −30 40] (r = 0.665) and [32 16 −12] (r = 0.992), indicating that the functional and anatomical terms associated with these SBC maps closely resemble those identified in PTSD. By contrast, a more moderate correlation was found for the SBC map seeded at [−6 48 −6] (r = 0.380). Notably, the SBC map seeded in the 4th cluster [−50 −20 12] identified in the ChP metaanalysis yielded terms primarily linked to motor processing (“motor”, “movement”, “SMA”).

**Table 4:** Nimare decoding results (expressed as correlation coefficient) of the SBC maps—centered on clusters of reduced regional grey matter volume from the ChP and PTSD meta-analyses and the conjunction analysis. All reported coordinates are expressed in the MNI standard space. Abbreviations: ChP, chronic pain; PTSD, post-traumatic stress disorder; Conj, conjunction analysis; n.c., not computable.

|  | Conj | ChP dataset |  |  |  | PTSD dataset |
| --- | --- | --- | --- | --- | --- | --- |
| Terms | [-8 48 -2] | [2 -30 40] | [-6 48 -6] | [32 16 -12] | [-50 -20 12] | [2 34 10] |
| affective | 0.686 | 0.494 | 0.684 | 0.391 | - | - |
| amygdala hippocampus | 0.754 | 0.788 | 0.724 | 0.688 | - | 0.786 |
| autonomic | 0.754 | 0.788 | 0.724 | 0.688 | - | 0.786 |
| face | 0.754 | 0.788 | 0.724 | 0.688 | - | 0.786 |
| limbic | 0.638 | 0.451 | 0.61 | 0.545 | - | 0.567 |
| memory | 0.670 | 0.730 | 0.668 | 0.559 | - | 0.586 |
| ofc | 0.754 | 0.788 | 0.724 | 0.688 | - | 0.786 |
| reward | 0.788 | 0.766 | 0.791 | 0.584 | - | 0.584 |
| semantic | 0.754 | 0.788 | 0.724 | 0.688 | - | 0.786 |
| striatum | 0.754 | 0.788 | 0.724 | 0.688 | - | 0.786 |
| detected | - | - | - | - | 0.495 | - |
| motor | - | - | - | - | 0.738 | - |
| movement | - | - | - | - | 0.589 | - |
| primary motor | - | - | - | - | 0.589 | - |
| reading | - | - | - | - | 0.589 | - |
| reflecting | - | - | - | - | 0.648 | - |
| sma | - | - | - | - | 0.495 | - |
| speech production | - | - | - | - | 0.589 | - |
| ChP-PTSD term profile correlation (r) |  | 0.665 | 0.380 | 0.992 | n.c. |  |

**Figure 6.**
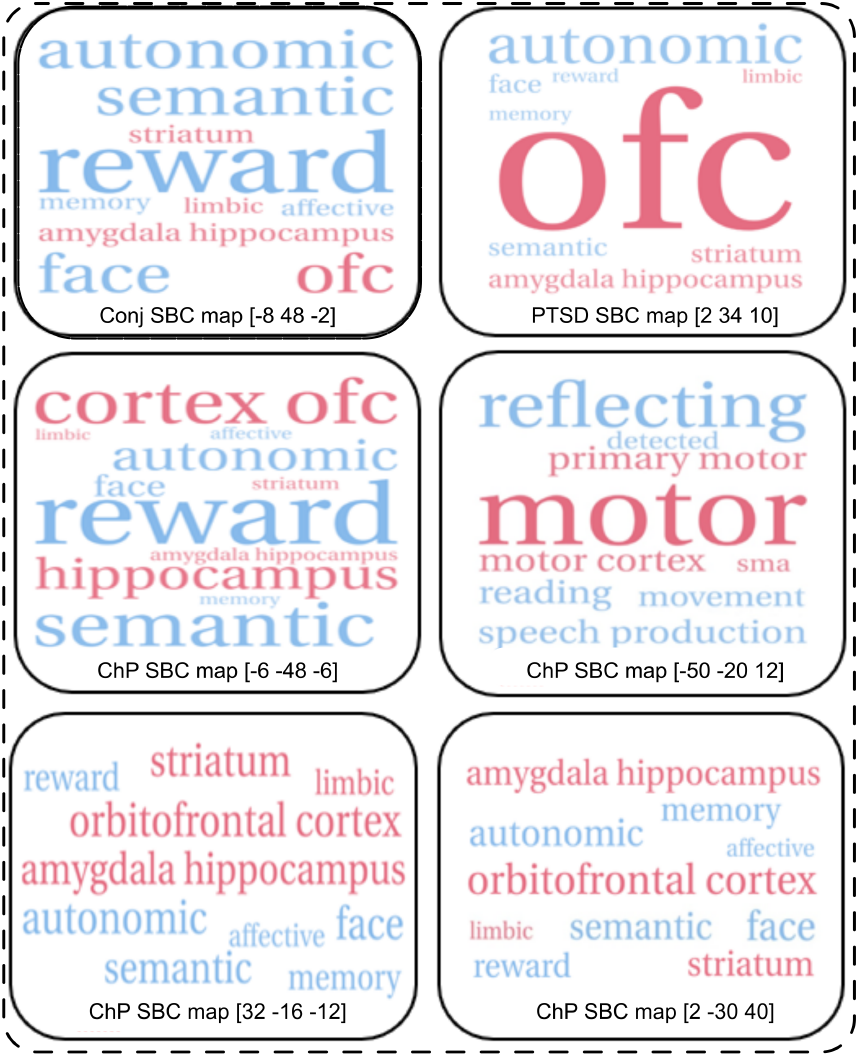
Word clouds of terms obtained from NiMARE decoding of the SBC maps—seeded at the peak coordinate (in brackets) of reduced regional grey matter volume from the ChP and PTSD meta-analyses and the conjunction analysis. Terms represent cognitive processes (blue) and anatomical regions (pink). Font size is proportional to the absolute correlation coefficient (r) between each term and the decoded SBC map. Abbreviations: ChP, chronic pain; PTSD, post-traumatic stress disorder; Conj, conjunction; SBC, seed-based functional connectivity.

### 3.6. Seed-based resting-state functional connectivity maps: neurotransmitter receptors/transporters profiling

See Figure 7 and Tables 5 for the results of this section. The spatial correlation analysis between the SBC map obtained from the Conj analysis—i.e., the region of converging grey matter volume reductions across the ChP and PTSD datasets—and the selected neurotransmitter receptor/transporter distribution maps revealed significant correlations with the dopaminergic (FDOPA map), serotoninergic (SERT maps) and opioid system (MOR maps). Unexpectedly, no spatial correlation was found between the SBC maps seeded in the clusters obtained from both ChP and PTSD meta-analyses.

**Table 5:** Spatial correlation analysis between the SBC map obtained from the Conj analysis and the ChP and PTSD meta-analyses and the selected neurotransmitter receptor/transporter distribution maps. All reported coordinates are expressed in the MNI standard space. * Significant results corrected for multiple comparisons (*p* < 0.05, Benjamini–Hochberg FDR-corrected). Abbreviations: Conj, conjunction; ChP, chronic pain; PTSD, post-traumatic stress disorder; SBC, seed-based connectivity.

| PET Map | Conj SBC map |  | ChP SBC maps |  |  |  |  |  |  |  | PTSD SBC map |  |
| --- | --- | --- | --- | --- | --- | --- | --- | --- | --- | --- | --- | --- |
|  | Seed in [-8 48 -2] |  | Seed in [2 -30 40] |  | Seed in [-6 48 -6] |  | Seed in [32 16 -12] |  | Seed in [-50 -20 12] |  | Seed in [2 34 10] |  |
|  | *p* | *Fisher's* *z* | *p* | *Fisher's* *z* | *p* | *Fisher's* *z* | *p* | *Fisher's* *z* | *p* | *Fisher's* *z* | *p* | *Fisher's* *z* |
| CB1 (Laurikainen et al., 2019)(Laurikainen et al., 2019) | 0.522 | 0.120 | 0.997 | 0.015 | 0.709 | 0.086 | 0.471 | 0.282 | 0.471 | 0.282 | 0.367 | 0.159 |
| CB1 (Normandin et al., 2015)(Normandin et al., 2015) | 0.054 | 0.330 | 0.997 | 0.147 | 0.149 | 0.288 | 0.471 | 0.159 | 0.471 | 0.159 | 0.367 | 0.146 |
| FDOPA (Garcia-Gomez et al., 2018)(Garcia-Larrea and Bastuji, 2018) | 0.013* | 0.321 | 0.997 | 0.071 | 0.149 | 0.194 | 0.834 | 0.043 | 0.834 | 0.043 | 0.078 | 0.252 |
| KOR (Vijay et al., 2018)(Vijay et al., 2018) | 0.066 | 0.255 | 0.997 | 0.239 | 0.149 | 0.225 | 0.471 | 0.234 | 0.471 | 0.234 | 0.147 | 0.308 |
| KappaOp (Shokri-Kojori et al., 2022)(Shokri-Kojori et al., 2022) | 0.522 | 0.097 | 0.997 | 0.001 | 0.771 | 0.042 | 0.471 | 0.312 | 0.471 | 0.312 | 0.367 | 0.158 |
| MOR (Kantonen et al., 2020)(Kantonen et al., 2020) | 0.013* | 0.468 | 0.997 | 0.029 | 0.149 | 0.325 | 0.513 | 0.201 | 0.513 | 0.201 | 0.323 | 0.277 |
| MOR (Turtonen et al., 2021)(Turtonen et al., 2021) | 0.042* | 0.486 | 0.997 | 0.043 | 0.149 | 0.345 | 0.471 | 0.263 | 0.471 | 0.263 | 0.323 | 0.314 |
| MU (Kantonen et al., 2020)(Kantonen et al., 2020) | 0.058 | 0.381 | 0.997 | -0.036 | 0.225 | 0.251 | 0.471 | 0.210 | 0.471 | 0.210 | 0.367 | 0.209 |
| MU (Turtonen et al., 2021)(Turtonen et al., 2021) | 0.058 | 0.405 | 0.997 | -0.037 | 0.225 | 0.265 | 0.471 | 0.272 | 0.471 | 0.272 | 0.367 | 0.244 |
| NAT (Hesse et al., 2017)(Hesse et al., 2017) | 0.522 | -0.057 | 0.997 | 0.100 | 0.709 | -0.038 | 0.837 | -0.033 | 0.837 | -0.033 | 0.190 | 0.168 |
| SERT (Savli et al., 2012)(Savli et al., 2012) | 0.013* | 0.278 | 0.997 | -0.021 | 0.217 | 0.141 | 0.842 | -0.025 | 0.842 | -0.025 | 0.297 | 0.130 |
| SERT (Fazio et al., 2016)(Fazio et al., 2016) | 0.013* | 0.278 | 0.997 | -0.024 | 0.149 | 0.166 | 0.842 | -0.018 | 0.842 | -0.018 | 0.130 | 0.223 |
| SERT (Beliveau et al., 2017)(Beliveau et al., 2017) | 0.086 | 0.166 | 0.997 | -0.050 | 0.531 | 0.076 | 0.834 | -0.047 | 0.834 | -0.047 | 0.182 | 0.173 |

**Figure 7.**
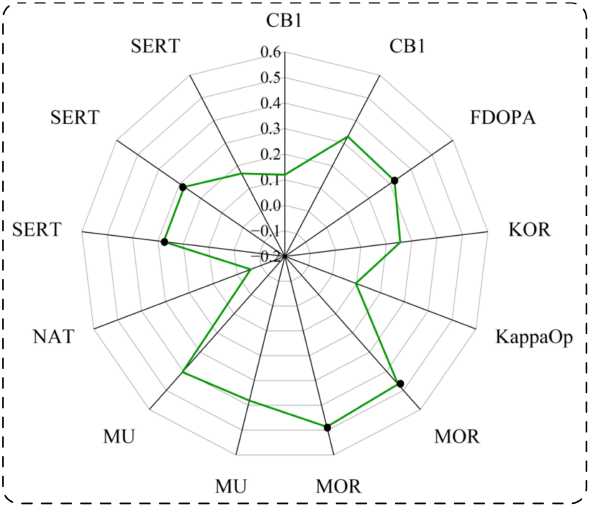
Radar charts showing the spatial correlations between selected PET maps and the SBC map seeded at the peak coordinate resulting from the conjunction analysis. Black dots indicate significant correlations (*p* < 0.05 FDR-corrected). Abbreviations: Conj, conjunction analysis; SBC, seed-based functional connectivity; FDOPA, 18F-DOPA; KOR/KappaOp, *κ*-opioid receptor; MOR and MU, *µ*-opioid receptor; NAT, noradrenaline transporter; SERT, serotonin transporter; CB1, cannabinoid receptor 1.

## 4. Discussion

This pre-registered transdiagnostic meta-analysis aimed to determine whether the morphological brain abnormalities observed in ChP and PTSD — a prototypical stress-related condition — show any degree of convergence, despite their distinct pathophysiological profiles and behavioral symptoms. To this aim, we examined volumetric alterations in two independent meta-analytic datasets — one comprising studies on ChP (records, n = 60) and the other on PTSD (records, n = 20) - to identify both similarities and disorder-specific patterns of anatomical alteration. We further complemented the meta-analytic findings with an unbiased characterization of the identified regions of abnormalities, embedding the structural findings into functional networks, cognitive domains, and neurotransmitter systems. Although the meta-analytic results (“ChP meta-analysis” and “PTSD meta-analysis”) initially revealed widespread and robust structural alterations in each disorder separately—confirming the validity of our analytical approach—they also highlighted a focal and specific overlap: reduced gray matter volume in the medial prefrontal/anterior cingulate cortex. Beyond this key overlapping region, a transdiagnostic reappraisal uncovered a more integrated picture: the patterns of volume loss in both conditions were embedded within a shared architecture of large-scale circuits, mapping onto the mesocorticolimbic/reward network as well as the DMN and the salience network, with a prominent role for the mesocorticolimbic/reward network. Decoding analyses supported these findings by showing that these large-scale circuits are implicated in highly similar processes —including autonomic regulation, reward processing, and affective functions— and are tightly anchored in mesocorticolimbic structures.

In what follows, we outline several main findings whose collective interpretation points to shared neural substrates between ChP and PTSD, thereby supporting the view of ChP as a stress-related disorder.

Firstly, the results of the meta-analyses conducted separately for each disorder (“ChP meta-analysis” and “PTSD meta-analysis”) indicated partially distinct patterns of gray matter volume reduction relative to control subjects. Specifically, the ChP dataset revealed 4 clusters of reduced volume located primarily within the ventromedial prefrontal cortex (corresponding mainly to the medial frontal orbital gyrus), as well as within the middle cingulate cortex. Additional clusters involved the anterior and posterior portions of the bilateral insula, extending into adjacent regions. In contrast, the PTSD dataset revealed a single prominent cluster of volume reduction centered mainly in the anterior cingulate cortex and extending to the dorsomedial prefrontal cortex. Importantly, these effects were not confined to the peak locations: in both conditions, the clusters extended into surrounding medial prefrontal and cingulate regions, resulting in partial spatial overlap despite distinct peaks. Indeed, the conjunction analysis demonstrated that ChP and PTSD share a common region of reduced gray matter volume within the medial prefrontal/anterior cingulate cortex.

Secondly, the comparison of the two datasets (i.e, metaregression) revealed that PTSD patients with respect to ChP presented volume loss within the anterior cingulate cortex and dorsomedial prefrontal cortex—regions critically involved in emotion regulation, traumatic memory integration, and threat appraisal (Zhou et al., 2025). This finding likely reflects PTSD-specific deficits: impaired topdown control over hyperarousal and intrusive traumatic memories, as well as compromised ability to modulate emotional responses to threat. In contrast, the ChP dataset did not show any specific volume reductions relative to PTSD. While on one side, this disorder difference is consistent with a pronounced anterior cingulate and dorsomedial prefrontal cortices compromise in PTSD, on the other, it suggests that the structural abnormalities observed in ChP fall within the PTSD profile. Although the number of studies included in the two meta-analyses was unbalanced, with substantially more records available for ChP than for PTSD, this difference would more likely reduce sensitivity to detect effects in PTSD rather than inflate them and may have resulted in a conservative outcome in the PTSD < ChP contrast. In agreement, the pooled meta-analysis (including data from both ChP and PTSD studies) revealed extensive clusters of volume reduction—including the anterior cingulate cortex, bilateral medial prefrontal cortices, bilateral insula, and subcortical structures—with low-to-moderate heterogeneity (*I*^2^ = 6–32%), generally supporting a shared neurobiological substrate in particular in the cluster comprising the anterior region of the left insula cortex.

Thirdly, after generating the functional connectivity maps (i.e., SBC maps) anchored at peaks of gray matter volume reduction—using a subset of 7T resting-state fMRI data from healthy participants in the HCP—formal spatial similarity analyses indicated that both disorders converged on the mesocorticolimbic/reward network, as well as on the DMN and salience network, with a prominent role of the reward-related circuitry. In ChP, the mesocorticolimbic/reward network emerged as the most consistently involved system, being associated with 3 out of 4 SBC maps and showing stable empirical overlap across seeds. Notably, although PTSD was characterized by a single disorder-specific SBC map, the mesocorticolimbic/reward network showed the highest empirical overlap (emp = 0.743) among all canonical networks, indicating a particularly strong coupling with reward-related circuitry. These robust functional convergences across the two disorders support the hypothesis of a shared, stress-related, systems-level signature. Importantly, and in line with the distinct neurofunctional profiles of the two conditions, the functional connectivity patterns revealed differential engagement of sensory-related systems. Specifically, the ChP functional maps converged onto the sensorimotor (with the dorsal-attention and frontoparietal networks, whereas the PTSD maps converged onto the visual network.

Fourthly, the decoding of the functional connectivity maps revealed a substantial overlap in the associated cognitive terms, with moderate to very high correlations between the profile derived from the PTSD cluster of volume loss and those obtained from 3 of the 4 ChP clusters. These overlapping profiles were robustly associated with autonomic, reward, and affective domains, and mesolimbic structures.

Fifthly, only the functional connectivity map anchored in the shared cluster of volume loss (obtained from the conjunction analysis and located in the frontal medial orbital gyrus/anterior cingulate cortex) exhibited a coherent neurotransmitter fingerprint, showing significant spatial correspondence with the PET-derived maps for FDOPA, opioid, endocannabinoid, and serotonergic receptors. This selective PET correspondence in the conjunction-seeded network suggests that this shared hub concentrates neuromodulatory architecture relevant to stress–emotion–reward–pain axis, whereas disorder-specific seeds likely exhibit more heterogeneous receptor co-localization; the lack of significance elsewhere should be viewed as limited sensitivity rather than evidence of absence.

Despite the high comorbidity between ChP and PTSD, previous neuroimaging research has remained largely disorder-specific, with several disorder-specific metaanalyses providing insights into their respective volumetric alterations. However, while meta-analytic findings on PTSD appear relatively reproducible across studies and generally consistent with our results, those on ChP remain rather inconsistent, both across investigations and with respect to our findings. Consistent with the findings from our PTSD dataset, the two most comprehensive and recent meta-analyses to date (Xiao et al., 2022; Pankey et al., 2022) converged in identifying gray matter volume reductions in medial prefrontal regions. In addition, our observation that PTSD-related volumetric reductions are primarily anchored within the DMN, salience, visual, and mesocorticolimbic/reward networks closely aligns with the network-level involvement reported by Pankey et al. (2022).

With respect to ChP, the largest meta-analysis to date (Henn et al., 2023) (records, n = 75) reported structural gray matter reductions in patients within the anterior cingulo-insular and medial prefrontal regions. Among these regions, the right anterior insula was the only cluster consistently identified across both this study and the present meta-analysis. Another recent meta-analysis, conducted within a transdiagnostic framework encompassing not only ChP but also depression and anxiety disorders (Brandl et al., 2022) (records, n = 65), found evidence of increased gray matter volumes in several subcortical regions, with the cerebellum, along with decreased volumes in the right anterior insula. To functionally contextualize these structural findings, the authors projected each significant cluster onto a canonical functional network parcellation and showed that the grey matter volume increases were mainly located within the limbic network and DMN, whereas the volume decreases fell predominantly within the salience and frontoparietal networks. In this case as well, we replicated only the right anterior insula findings; nevertheless, our overall functional network profile partially converged with their observation of limbic, DMN, salience, and frontoparietal networks involvement. Finally, the most recent meta-analytic study (Zeng et al., 2025) (records, n = 47) reported decreased gray-matter volumes in several regions encompassing the posterior portions of the bilateral insula and the rolandic operculum, as well as the left superior frontal gyrus extending into the anterior and mid-cingulate cortices. In this comparison, our findings replicated only the gray-matter reductions in the left posterior insular cortex. The discrepancies between our findings and previous meta-analyses (Henn et al., 2023; Brandl et al., 2022; Zeng et al., 2025) can be attributed to both methodological differences and the greater heterogeneity of the populations considered in those studies. For example, studies included in those meta-analyses often featured more diverse participant groups, such as individuals outside the age range of 18 to 65 years or those with conditions like episodic migraine or systemic disorders that can independently affect brain morphology. Additionally, we focused on studies employing VBM and stricter methodological thresholds, while some previous works included studies with fewer than 10 participants per group or employing less rigorous statistical methods. These decisions were made to ensure a more homogeneous dataset, which we believe contributes to the robustness of our results. The consistency of our meta-analytic results in the ChP dataset is supported by the fact that the regions showing reduced cortical volume (medial prefrontal-cingular regions, precuneus, and anterior and posterior insula cortices) are key-hubs of functional networks-namely the mesocorticolimbic/reward circuitry, DMN, salience, frontoparietal, and sensorimotor/dorsal-attention networks-that have been widely implicated in ChP pathophysiology. In particular, alterations in the mesocorticolimbic/reward circuitry appear central in ChP (Baliki et al., 2011, 2012; Vachon-Presseau et al., 2016a; Martucci et al., 2018; Borsook et al., 2016), possibly reflecting dysregulation of reward valuation and motivational learning processes in the context of persistent pain. Additionally, aberrant DMN activity is associated with maladaptive self-referential processes (van Ettinger-Veenstra et al., 2019; Otti et al., 2013; Kucyi et al., 2014) and fluctuations in perceived pain intensity via its coupling with the salience network (Jahn et al., 2021).

Shifting from a shared to a disorder-specific perspective, functional connectivity maps centered on regions of reduced volume revealed that both ChP and PTSD exhibit network-level patterns encompassing 3 common large-scale systems—namely, the mesocorticolimbic/reward, DMN, and salience networks. Notably, the mesocorticolimbic/reward network emerged as a key circuit of convergence in both conditions. In ChP, it was anchored in 3 out of 4 clusters showing abnormal volumetry, while it consistently co-occurred with the DMN alone, with the DMN and frontoparietal networks, and with the salience and frontoparietal networks. In PTSD, by contrast, it co-occurred with the DMN, salience, and visual networks, showing the highest empirical overlap across all considered networks (emp = 0.743). This general pattern suggests that a vulnerability within the mesocorticolimbic/reward circuit may represent a shared mechanism through which chronic stress reshapes large-scale network dynamics in both ChP and PTSD. This interpretation is consistent with evidence showing that the mesocorticolimbic/reward circuit is a major target of stress, as reflected by the pronounced stress-induced dysregulation of its dopaminergic signaling (Baik, 2020; Cabib and Puglisi-Allegra, 1996), and by its central role in modulating stress responses (Vachon-Presseau, 2018). In this view, an initial stress-induced imbalance within a susceptible mesocorticolimbic/reward system may trigger abnormal activity of the salience network, which in turn disrupts the DMN. The mesolimbic dopamine neurons (Schultz, 1997) — originating in the ventral tegmental area and projecting to the nucleus accumbens and prefrontal cortex — and the salience network (Seeley et al., 2007; Craig and Craig, 2009; Menon and Uddin, 2010) are both involved in detecting behaviorally relevant stimuli and in reorienting behavior. Accordingly, they have been shown to robustly interact. Converging animal studies have shown that mesolimbic dopamine neurons may modulate the activity of salience network hubs (Roelofs et al., 2017; Helbing et al., 2016), while seminal human studies have demonstrated that elevated dopamine synthesis is associated with increased functional connectivity within the salience network (McCutcheon et al., 2019). Finally, in ChP, the nucleus accumbens shows reduced low-frequency fluctuations and altered coupling with the anterior cingulate cortex, a core hub of the salience network (Makary et al., 2020). Modulating the dynamic balance between internally directed (DMN) and externally oriented (frontoparietal) systems, salience network dysregulation can propagate to self-referential and cognitive processing domains, as observed in both PTSD and ChP. These cascading alterations could represent the network pathway through which a dopaminergic vulnerability evolves into PTSD or ChP, while symptom expression diverges according to the dominant sensory pathways through which the stressor is processed—visual and threat-related in PTSD versus interoceptive and somatosensory in ChP. Interestingly, in PTSD, the visual network emerged alongside the mesocorticolimbic/reward, DMN, and salience networks, suggesting enhanced visuo-emotional integration — a configuration consistent with hypervigilant threat monitoring and intrusive re-experiencing phenomena. In contrast, in ChP, we observed the additional involvement of the sensorimotor and dorsal attention networks. Although this configuration might appear to represent a purely sensory domain, it more likely reflects more a perceptual–attentional component. The co-activation of sensorimotor and dorsalattention systems suggests sustained monitoring of bodily and nociceptive signals — a process of somatic hypervigilance rather than mere sensory encoding. Notably, the selective involvement of the frontoparietal network in ChP, not observed in PTSD, may represent a secondary consequence of prolonged stress-related dysregulation within the reward-salience-DMN axis, emerging only after sustained exposure to interoceptive or nociceptive overload. In this view, executive control systems become progres-sively compromised as they attempt to regulate persistent stress signals. In PTSD, such alterations may remain less pronounced, as the disorder often follows an episodic course and can show substantial functional recovery following effective treatment, possibly preventing the long-term neurofunctional deterioration observed in ChP conditions.

Interestingly, our findings revealed a shared volume reduction within a region encompassing the medial prefrontal/anterior cingulate cortex, whose underlying functional network mapped onto both the mesocorticolimbic/reward circuitry and the DMN. Notably, this region showed robust spatial correspondence with multiple neuromodulatory systems-dopaminergic, opioidergic, endocannabinoid, and serotonergic—suggesting that this shared hub integrates neurochemical mechanisms central to reward (Bromberg-Martin et al., 2010), affective regulation (Dayan and Huys, 2009), pain processing (Zubieta et al., 2001), and stress responsivity (Hillard et al., 2017). This common region of volume reduction may therefore represent a critical node in neural dysregulations that mediate the impact of chronic stress on different conditions, including PTSD and ChP. This region may as such, present a novel target for interventions that target this region and related functions such as emotion regulation or reward-based processing, including angiotensin II blockade(Zhou et al., 2023; Xu et al., 2023; Zhou et al., 2019) or real-time neurofeedback targeting this core node and enhancing motivation and reward processing(Aupperle et al., 2024; Zhang et al., 2025) It is important to note, however, that the specific localization of the observed spatial convergence should be interpreted with caution. The relatively large smoothing kernel inherent to SDM and inter-study variability can contribute to apparent overlap between anatomically adjacent regions. Nonetheless, the persistence of overlapping clusters within the medial prefrontal–cingulate territory—regions consistently implicated in stress regulation and affective control, and the high spatial correspondence with dopaminergic, serotonergic, opioid, and endocannabinoid PET density maps suggests that the convergence may reflect a genuine shared structural vulnerability consistent with stress- and reward/salience-related mechanisms implicated in both conditions.

### Limitations

In interpreting our results, several limitations should be considered. First, the number of studies included for PTSD (n = 20) was markedly smaller than for ChP (n = 60), which could theoretically bias the pooled analyses by assigning greater statistical weight to the larger dataset. However, this imbalance is unlikely to have inflated the observed convergence; if anything, it may have led to a more conservative estimate of shared effects, as regions identified as common had to emerge robustly in both conditions despite the lower statistical power of the PTSD dataset.

Second, few studies included in the present work systematically reported comorbidity data, particularly with respect to ChP conditions in PTSD samples and PTSD diagnoses in ChP samples. This lack of information represents a potential confounding factor, as unreported comorbidities may contribute to an apparent neurobiological overlap between the two disorders. In addition, depression represents a major source of potential confounding, given its high prevalence as a comorbid condition in both PTSD and ChP populations(Flory and Yehuda, 2015; Roughan et al., 2021; Bair et al., 2003). As a consequence, some of the observed structural alterations may partly reflect the neurobiological correlates of depression rather than disorder-specific changes. However, it is worth noting that the abnormal anatomical regions identified in the two separate meta-analyses were largely distinct. In particular, the anterior cingulate cortex emerged as a significant cluster in the PTSD meta-analysis and in the PTSD < ChP contrast, while no significant clusters were observed in the ChP < PTSD contrast. Although comorbidity rates could not be formally assessed, this pattern may tentatively suggest limited cross-contamination between the diagnostic groups included in the present analyses.

Third, we identified the functional networks associated with regions of reduced gray matter volume by employing resting-state functional connectivity data from a subset of healthy participants of the HCP—one of the most comprehensive and methodologically rigorous neuroimaging datasets currently available. This choice provided a high-quality normative reference for inferring the potential large-scale network engaged in specific disorders. Rather than a limitation, this methodological decision represents a major strength of the present study. Large-scale functional architectures are not only stable across individuals, physiological states, and clinical conditions, but are also evolutionarily conserved across mammalian species (Gozzi and Schwarz, 2016; Vincent et al., 2007), supporting the notion that connectivity patterns capture fundamental organizational principles of the brain. Therefore, mapping the connectivity profiles of structurally altered regions within such a rigorous normative framework allows for the reliable identification of systems likely to be involved or compromised, offering a principled and generalizable way to link structural abnormalities to stable, population-defined functional organization. Consistently, the convergence between the connectivity profiles in ChP and PTSD and the large-scale networks previously reported to be disrupted in these conditions further supports the validity of our multimodal approach.

An additional limitation concerns differences in pharmacological exposure across diagnostic groups. PTSD samples were largely medication-free, whereas ChP samples showed substantial heterogeneity in medication use (see Tables S4, S2), including analgesic, psychotropic, and anti-inflammatory agents. As medication use was not systematically controlled, its potential influence on brain morphometric findings cannot be entirely excluded. Importantly, however, the observation of overlapping alterations within core large-scale networks (i.e., mesocorti-colimbic circuitry, DMN, and salience network) emerged despite this marked asymmetry in pharmacological exposure. Given that PTSD samples were predominantly medication-free, the presence of shared neuroanatomical patterns across the two conditions is unlikely to be fully attributable to medication effects alone, and instead tentatively supports the existence of partially shared neurobiological mechanisms underlying PTSD and ChP.

Another limitation concerns the inability to account for neuroinflammatory mechanisms. Pro-inflammatory cytokines, including IL-6, have been implicated in the pathophysiology of both PTSD and ChP (Passos et al., 2015; Ji et al., 2018). However, inflammatory markers were not reported in the included studies, precluding any assessment of their potential contribution to the observed structural brain alterations.

## Conclusion

Our transdiagnostic meta-analytic approach, complemented by an extended profiling of the underlying functional networks, indicates that the patterns of gray matter volume loss observed in ChP and PTSD—although largely distinct—are anchored within a shared core of large-scale networks, consistently mapping onto the mesocorticolimbic/reward system, the DMN, and the salience network. Among these, a particularly vulnerable mesocorticolimbic system might represent the most critical network through which stress responses can exert maladaptive effects leading to ChP and PTSD, with cascading influences on other large-scale systems involved in emotion regulation, salience processing, and executive control. By situating morphometric changes within broader network-level dynamics, this study supports the reconceptualization of ChP as a stress-related disorder while also highlighting potential pathways for targeted, network-based therapeutic interventions.

## Data Availability

All data produced in the present work are contained in the manuscript

## Data availability

The records selected for study are reported in the supplementary materials with the relevant details, while the coordinates of each selected study used for the metaanalyses are made available within the preregistration materials (osf.io/m2bzu).

## Supplementary Material

**Table S1:** Records selected for the chronic pain dataset with the relevant information. From each paper, we reported the source (Table, Main text or SM) from which the relevant coordinates were extracted. Abbreviations: CTRL: control subjects. Norm. space, normalization space; SM, supplementary materials; ns, not specified.

| CHRONIC PAIN DATASET |  |  |  |  |  |  |  |  |
| --- | --- | --- | --- | --- | --- | --- | --- | --- |
| Authors | TABLE | #Patients (Females) | Age M±SD | #CTRL (Females) | Age M±SD | Threshold employed to report the re-sults | Norm. space | Software |
| Apkarian et al. (2004) | In text | 26(16) | 42.9±ns | 26(16) | 42.9±ns | Permutation-based cluster-level corrected | MNI | SPM99 |
| Baliki et al. (2011) | Table S1 | 36(13) | 48.2±11.38 | 46(26) | 38.77±12.5 | p<0.01 TFCE-corrected | MNI | FSL 4 |
| Baliki et al. (2011) | Table S1 | 28(24) | 40.57±7.4 | 46(26) | 38.77±12.5 | p<0.01 TFCE-corrected | MNI | FSL 4 |
| Baliki et al. (2011) | Table S1 | 20(20) | 53.5±7.4 | 46(26) | 38.77±12.5 | p<0.01 TFCE-corrected | MNI | FSL 4 |
| Barad et al. (2014) | Table A1 | 15(15) | 44±ns | 15(15) | 44.1±ns | p<N.R. FDR-corrected (corresponding, p=0.0005) | MNI | SPM8 |
| Bishop et al. (2018) | Table 4 | 74(60) | 45.7±13.03 | 31(18) | 44.13±13.15 | p<= 0.05 TFCE-corrected | MNI | FSL |
| Boehme et al. (2020) | Table 2 | 31(31) | 39.2±11.4 | 29(29) | 42.7±10.1 | p<0.001 | MNI | SPM12 |
| Ceko et al. (2013) | Table 2 | 14(14) | 55±2.9 | 13(13) | 55.4±3.7 | p<0.05 RFT-corrected cluster-level; p<0.01 voxel-wise | MNI | SPM8 |
| Ceko et al. (2013) | Table 2 | 14(14) | 42.4±5.9 | 15(15) | 43.1±5.3 | p<0.05 RFT-corrected cluster-level; p<0.01 voxel-wise | MNI | SPM8 |
| Coppola et al. (2017) | Table 2 | 20(14) | 31.1±10.2 | 20(13) | 28.5±4.1 | p<0.001 | MNI | SPM12 |
| Domin et al. (2021) | In Text | 24(20) | 50.7±14 | 33(19) | 54.42±13.49 | p<0.05 FWE-corrected voxel-wise | MNI | SPM12 |
| Arkin et al. (2017) | Table 3 | 23(4) | 48±9 | 48(30) | 47±9 | p<0.001 | MNI | SPM8 |
| Fallon et al. (2013) | Table 1 & 2 | 16(16) | 38.5±8.45 | 15(15) | 39.4±8.7 | p< 0.05 FWE-corrected cluster-level; p<0.001 voxel-wise | MNI | SPM8 |
| Gandola et al. (2017) | In text | 35(27) | 60.1±9.4 | 35(27) | 57.9±9.9 | p< 0.05 FWE-corrected cluster-level; p<0.001 voxel-wise | MNI | SPM8 |
| Geha et al. (2008) | In text | 22(19) | 40.7±2.3 | 22(19) | 40.5±2.3 | p<0.05 cluster-level corrected (permutation-based) | MNI | FSL 4 |
| Gustin et al. (2011) | Table 3 | 21(17) | 54.7±2.1 | 30(24) | 53.6±3.2 | p<0.01 FDR-corrected voxel-wise | MNI | SPM5 |
| Gwilym et al. (2010) | Table 1 | 16(8) | 68±ns | 16(8) | ns±ns | p<0.001 | MNI | FSL |
| Ikeda et al. (2018) | Table 2 | 23(15) | 47.7±14.1 | 17(9) | 47.4±14.1 | p<0.001 | MNI | SPM12 |
| Ivo et al. (2013) | Table 2 | 14(8) | 54±ns | 14(ns) | ns±ns | p<0.001 | MNI | SPM8 |
| Kairys et al. (2015) | Table 1 | 33(33) | 39.5±12 | 33(33) | 39±11.6 | p<0.001 | MNI | SPM8 |
| Kuchinad et al. (2007) | Table 1 | 10(10) | 52±ns | 10(10) | 45±ns | t>3.0 (p=0.007) -reported only p<0.005 | TAL | In-house pipeline |
| Lai et al. (2016) | Table 2 | 33(27) | 39.7±10.7 | 33(27) | 39.7±11.1 | p<0.05 FWE-corrected cluster-level; p<0.005 voxel-wise | MNI | SPM8 |
| Lee et al. (2019) | Table 1 | 12(ns) | ns±ns | 14(ns) | ns±ns | p<0.05 FWE-corrected voxel-wise | MNI | SPM12 |
| Liao et al. (2018) | Figure 2 | 30(26) | 56.5±6.8 | 30(26) | 55.2±5.7 | p<0.05 FWE-corrected cluster-level (permutation-based) | MNI | FSL 5 |
| Li et al. (2017) | Table 2 | 28(13) | 45.86±11.17 | 28(13) | 44.89±7.67 | p<0.05 FWE-corrected voxel-wise | MNI | SPM8 |
| Li et al. (2018b) | Table 3 | 16(4) | 41.6±13.6 | 16(0) | 31.1±11 | p<0.001 | MNI | SPM12 |
| Liu et al. (2022) | Table 2 | 34(18) | 53.06±10.91 | 29(14) | 54.21±6.33 | p<0.05 FDR-corrected | MNI | SPM12 |
| Luchtmann et al. (2014) | Table 1 | 12(ns) | 43.9±12.9 | 12(ns) | ns±ns | p<0.0001 | MNI | SPM8 |
| Maeda et al. (2013) | Table 1 | 28(20) | 48.1±9.6 | 28(17) | 47.3±9.9 | p<0.05 FDR | MNI | SPM8 |
| Martikainen et al. (2013) | In text | 16(8) | 38±11 | 16(8) | 36±11 | p<0.001 uncorr | MNI | SPM8 |
| Mordasini et al. (2012) | Table 1 | 20(0) | 40±14 | 20(0) | 43±19 | p<0.001 uncorr | TAL | SPM5 |
| Naegel et al. (2014) | Table S1 | 23(7) | 47.96±10.56 | 78(22) | 42.78±11.44 | p<0.001 uncorr | MNI | SPM8 |
| Neeb et al. (2017) | Table 2 | 21(15) | 49.04±7.46 | 21(15) | 49.4±7.79 | p<0.001 uncorr | MNI | SPM8 |
| Neumann et al. (2023) | In text | 305(209) | 48.9±ns | 296(99) | 54.95±14.2 | p<0.05 FWE-corrected voxel-wise | MNI | SPM12 |
| Niddam et al. (2017) | Table 2 | 21(16) | 42.1±12.7 | 21(16) | 38±7.7 | p<0.005 uncorrected | TAL | SPM8 |
| Nimmman et al. (2022) | Table 4 | 108(11) | 49.8±6.5 | 62(8) | 49.1±7.4 | p<0.05 FWE-corrected cluster-level; p<0.001 voxel-wise | MNI | SPM12 |
| Obermann et al. (2013) | Table 2 | 60(36) | 62±13.2 | 49(28) | 61.8±9 | p<0.001 | MNI | SPM8 |
| Öhlmann et al. (2021) | Table 2 | 23(23) | 46.9±10.92 | 23(23) | 43.74±12.17 | p<0.05 TFCE-corrected | MNI | SPM12 |
| Pleger et al. (2014) | Table 2 | 20(11) | 41.8±9.8 | 20(11) | 41.6±9.6 | p<0.05 FWE-corrected cluster-level; p<0.001 voxel-wise | MNI | SPM8 |
| Pomares et al. (2017) | Table 2a | 26(26) | 61±5.4 | 25(25) | 61±7.6 | z=2.3 ( p<0.02) -reported fino 3.09 (p=0.001) | MNI | SPM8 |
| Riederer et al. (2017) | Table 5 | 23(15) | 43.1±10.5 | 29(20) | 42.4±9.8 | p<0.001 uncor | MNI | SPM8 |
| Schelliga et al. (2024) | Table 2 | 26(15) | 45.3±10.77 | 25(13) | 41.04±18.53 | p<0.001 | MNI | SPM12 |
| Schweinhardt et al. (2008) | Table 3 | 14(14) | 25.7±5.1 | 14(14) | 25.6±6 | p<0.05 FWE-corrected cluster-level; voxel-wise p<0.01 | MNI | In-house pipeline |
| Schmidt-Wilcke et al. (2010) | Table 1 | 11(9) | 52.2±8.9 | 11(9) | 51.3±8.6 | p<0.001 | MNI | SPM5 |
| Schmidt-Wilcke et al. (2006) | Table 2 | 18(9) | 50.4±6.8 | 18(9) | 49.9±8.7 | p<0.001 | TAL | SPM99 |
| Schmidt-Wilcke et al. (2007) | Table 1 | 20(19) | 53.6±7.7 | 22(20) | 50.7±7.3 | p<0.001 | TAL | SPM2 |
| Shen et al. (2022) | Table 2 | 16(9) | 63.6±6.8 | 18(18) | 59.8±8 | p<0.005 TFCE-corrected | MNI | SPM12 |
| Shokouhi et al. (2018) | Table 2 | 12(10) | 51.1±12.7 | 16(10) | 44.4±11.6 | p<0.05 FWE-corrected cluster-level; voxel-wise | MNI | SPM8 |
| Sinding et al. (2016) | Table 1 | 12(7) | 59.4±12.1 | 13(10) | 59±3.4 | p<0.001 | MNI | SPM8 |
| Sundermann et al. (2019) | Table SM 1 | 25(25) | 52.6±11.1 | 21(21) | 52.24±9.79 | p<0.001 | MNI | SPM12 |
| Sundermann et al. (2019) | Table SM 1 | 23(23) | 54.1±12.36 | 21(21) | 52.24±9.79 | p<0.001 | MNI | SPM12 |
| Tan et al. (2019) | Table 2 | 26(21) | 52.1±8.81 | 27(25) | 51.11±5.42 | p<0.05 TFCE-corrected | MNI | SPM8 |
| Tang et al. (2021) | Table 2 | 25(11) | 59±9.84 | 21(9) | 58.48±10.34 | p<0.05 alphasim -as uncorrected | MNI | SPM8 |
| Tsai et al. (2018) | Table 2 | 36(20) | 58±7.7 | 19(15) | 55.6±8.2 | p<0.05 FWE-corrected cluster-level; p<0.05 voxel-wise threshold | MNI | SPM8 |
| Tsai et al. (2018) | Table 2 | 26(18) | 59±6.6 | 19(15) | 55.6±8.2 | p<0.05 FWE-corrected cluster-level; p<0.05 voxel-wise | MNI | SPM8 |
| Ung et al. (2014) | Figure 3 | 47(22) | 37.3±12.2 | 47(22) | 37.7±7.8 | p<0.001 | MNI | SPM8 |
| Valet et al. (2009) | Table 2 | 14(14) | 51.1±11.1 | 25(25) | 51.7±7.2 | p<0.05 FDR-corrected | MNI | SPM2 |
| van Velzen et al. (2016) | Table 2 | 19(19) | 48.1±11.6 | 19(19) | 49.4±14.3 | p<0.001 | MNI | FSL5 |
| Wang et al. (2023) | Table 2 & 3 | 28(2) | 30.6±ns | 29(2) | 29.55±ns | p< 0.05 FDR-corrected cluster-level; p<0.001 voxel-wise | MNI | SPM12 |
| Wang et al. (2019b) | Table 2 | 40(23) | 55.8±8.23 | 40(23) | 55.8±8.09 | p< 0.05 FDR cluster-level; p<0.001 voxel-wise | MNI | SPM12 |
| Wood et al. (2009) | Table 2 | 30(30) | 42±8.43 | 20(20) | 40.05±10.1 | t=3 (p<0.0043) | MNI | SPM2 |
| Wu et al. (2020) | Table 2 | 22(14) | 47.8±9.24 | 45(23) | 49.36±11.58 | p<0.001 | MNI | SPM12 |
| Yang et al. (2020) | Table 2 | 31(11) | 41.2±1.67 | 30(10) | 40.27±1.85 | p<0.001 | MNI | SPM8 |
| Younger et al. (2010) | Table 2 | 14(14) | 38±13.7 | 15(15) | ns±ns | p<0.05 FDR-corrected cluster-level; p<0.05 FDR-corrected voxel-wise | MNI | SPM8 |
| Zhang et al. (2018b) | Table 2 | 29(19) | 48.1±11.9 | 34(21) | 43.3±10.1 | p<0.05 FWE-corrected cluster-level; p<0.01 voxel-wise | MNI | SPM12 |

**Table S2:** Records included in the chronic pain dataset and their reported medication status, diagnosis, and assigned pathophysiological mechanisms.

| CHRONIC PAIN DATASET |  |  |  |
| --- | --- | --- | --- |
| Authors | Medication | Diagnosis | Pathophysiological mechanism |
| Apkarian et al. (2004) | not reported | chronic back pain | nociceptive |
| Baliki et al. (2010) | not reported | chronic back pain | nociceptive |
| Baliki et al. (2010) | not reported | complex regional pain syndrome | nociplastic |
| Baliki et al. (2010) | not reported | knee osteoarthritis | nociceptive |
| Barad et al. (2014) | opioids, anticonvulsants, antidepressants , NSAIDs , muscle relaxants | complex regional pain syndrome | nociplastic |
| Bishop et al. (2018) | medication-free | chronic musculoskeletal pain | nociceptive |
| Boehme et al. (2020) | medication-free | fibromyalgia | nociplastic |
| Ceko et al. (2013) | NSAIDs, antidepressants, muscle relaxants, anticonvulsants, cannabinoids, triptans | Fibromyalgia | nociplastic |
| Ceko et al. (2013) | NSAIDs, antidepressants, muscle relaxants, anticonvulsants, triptans | fibromyalgia | nociplastic |
| Coppola et al. (2017) | NSAIDs | chronic migraine | nociplastic |
| Domin et al. (2021) | NSAIDs, opioid, amitriptyline, pregabalin | complex regional pain syndrome | nociplastic |
| Arkink et al. (2017) | triptans, ergotamine, indomethacin, lithium, verapamil, oxygen | chronic cluster headache | nociplastic |
| Fallon et al. (2013) | pregabalin, gabapentin, amitriptyline, analgesics | fibromyalgia | nociplastic |
| Gandola et al. (2017) | not reported | osteoarthritis (rhizarthrosis) | nociceptive |
| Geha et al. (2008) | not reported | chronic complex regional pain syndrome (CRPS) | nociplastic |
| Gustin et al. (2011) | NSAIDs, opioid, amitriptyline, pregabalin | trigeminal neuropathic pain | neuropathic |
| Gwilym et al. (2010) | not reported | osteoarthritis | nociceptive |
| Ikeda et al. (2018) | anticonvulsants, antidepressants, opioids, acetaminophen, NSAIDs, herbal medicines | Fibromyalgia, phantom pain, chronic low back pain | Not assigned |
| Ivo et al. (2013) | not reported | chronic low back pain | nociceptive |
| Kairys et al. (2015) | not reported | interstitial cystitis | nociplastic |
| Kuchinad et al. (2007) | not reported | fibromyalgia | nociplastic |
| Lai et al. (2016) | medication-free | chronic migraine without MOH | nociplastic |
| Lee et al. (2019) | not reported | burning mouth syndrome | neuropathic |
| Liao et al. (2018) | not reported | chronic knee osteoarthritis | nociceptive |
| Li et al. (2017) | not reported | trigeminal neuralgia | neuropathic |
| Li et al. (2018a) | not reported | chronic low back pain | nociceptive |
| Liu et al. (2022) | not reported | trigeminal neuralgia | neuropathic |
| Luchtmann et al. (2014) | opioids, NSAIDs | lumbar disc herniation | nociceptive |
| Maeda et al. (2013) | not reported | Carpal tunnel syndrome | neuropathic |
| Martikainen et al. (2013) | acetaminophen, NSAIDs, muscle relaxants, SSRIs, SNRIs, gabapentin | chronic back pain | nociceptive |
| Mordasini et al. (2012) | not reported | refractory chronic pelvic pain syndrome | nociplastic |
| Naegel et al. (2014) | not reported | chronic cluster headache | nociplastic |
| Neeb et al. (2017) | not reported | chronic migraine | nociplastic |
| Neumann et al. (2023) | not reported | chronic back pain, craniomandibular disorder | Not assigned |
| Niddam et al. (2017) | medication-free | chronic myofascial pain | nociplastic |
| Ninneman et al. (2022) | medication-free | veterans with widespread muskuloskeletal pain | nociplastic |
| Obermann et al. (2013) | antiepileptics | chronic trigeminal neuralgia | neuropathic |
| Öhlmann et al. (2021) | medication-free | chronic visceral pain | nociplastic |
| Pleger et al. (2014) | not reported | complex regional pain syndrome | nociplastic |
| Pomares et al. (2017) | not reported | Fibromyalgia | nociplastic |
| Riederer et al. (2017) | not reported | chronic pain in nondermatomal somatosensory deficits | nociplastic |
| Scheliga et al. (2024) | medication-free | small-fiber neuropathy | neuropathic |
| Schweinhardt et al. (2008) | medication-free | chronic vulvar pain | nociplastic |
| Schmidt-Wilcke et al. (2010) | not reported | persistent idiopathic facial pain | nociplastic |
| Schmidt-Wilcke et al. (2006) | analgesics | chronic back pain | nociceptive |
| Schmidt-Wilcke et al. (2007) | SSRIs, opioids | fibromyalgia | nociplastic |
| Shen et al. (2022) | carbamazepine, gabapentin | trigeminal neuralgia | neuropathic |
| Shokouhi et al. (2018) | not reported | Complex regional pain syndrome | nociplastic |
| Sinding et al. (2016) | not reported | burning mouth syndrome | neuropathic |
| Sundermann et al. (2019) | medication-free | Fibromyalgia | nociplastic |
| Sundermann et al. (2019) | medication-free | Osteoarthritis | nociceptive |
| Tan et al. (2019) | not reported | burning mouth syndrome | neuropathic |
| Tang et al. (2021) | not reported | postherpetic neuralgia | neuropathic |
| Tsai et al. (2018) | not reported | trigeminal neuralgia -right | neuropathic |
| Tsai et al. (2018) | not reported | trigeminal neuralgia -left | neuropathic |
| Ung et al. (2014) | medication-free | chronic low back pain | nociceptive |
| Valet et al. (2009) | medication-free | pain disorders | Not assigned |
| van Velzen et al. (2016) | not reported | complex regional pain syndrome | nociplastic |
| Wang et al.(2023)Wang et al. (2023) | not reported | Neuropathic pain | neuropathic |
| WangYuan et al.(2019)Wang et al. (2019a) | not reported | Primary trigeminal neuralgia | neuropathic |
| Wood et al.(2009)Wood et al. (2009) | not reported | Fibromyalgia | nociplastic |
| Wu et al.(2020)Wu et al. (2020) | carbamazepine | primary trigeminal neuralgia | neuropathic |
| Yang et al.(2020)Yang et al. (2020) | medication-free | chronic cervical spondylotic pain | nociceptive |
| Younger et al.(2010)Younger et al. (2010) | medication-free | Chronic myofascial temporomandibular pain | nociceptive |
| Zhang et al.(2018)Zhang et al. (2018b) | carbamazepine, gabapentin | trigeminal neuralgia | neuropathic |

**Table S3:** Records selected for the post-traumatic stress disorder (PTSD) dataset with the relevant information. From each paper, we reported the source (Table, Main text or SM) from which the relevant coordinates were extracted. * Traumatized-exposed controls (TECs) Abbreviations: CTRL: control subjects. Norm. space, normalization space; SM, supplementary materials; ns, not specified.

| PTSD DATASET |  |  |  |  |  |  |  |  |
| --- | --- | --- | --- | --- | --- | --- | --- | --- |
| Authors | TABLE | #Patients<br>(Females) | Age M±SD | #CTRL (Fe-<br>males) | Age<br>M±SD | Threshold employed | Norm.<br>space | Software |
| Bossini et al. (2017) | Table 2 | 19(9) | 40±9 | 19(4) | 41±6 | p<0.05 corrected -Montecarlo simulation | MNI | SPM8 |
| Chao et al. (2012) | Table 3 | 21(0) | 35.9±11.2 | 21*(0) | 35.2±13.1 | p<0.001 | MNI | SPM8 |
| Cheng et al. (2015) | Table 3 | 30(9) | 26.3±8.1 | 30(9) | 26.2±6.6 | p<0.05 FWE-corrected | MNI | SPM8 |
| Chen et al. (2009) | In text | 12(8) | 34.56±4.91 | 12*(8) | 33.25±5.27 | p<0.05 corrected (ns) | TAL | SPM2 |
| Chen et al. (2012) | Table 2 | 10(ns) | 40.8±6.8 | 20*(ns) | 34.3±5.6 | p<0.05 FWE-corrected | MNI | SPM8 |
| Corbo et al. (2005) | In text | 14(ns) | 33.36±12.06 | 14(ns) | 33.29±12.31 | p<0.05 FWE-corrected voxel-wise | MNI | In-house pipeline |
| Eckart et al. (2011) | In text | 20(ns) | 36.2±7.7 | 19*(ns) | 34.1±9.9 | p<0.005 | MNI | SPM5 |
| Herringa et al. (2012) | Table 2 | 13(2) | 28.9±4.2 | 15*(1) | 30.1±6.3 | p<0.001 | MNI | SPM8 |
| Kasai et al. (2008) | Table 2 | 18(0) | 52.8±3.4 | 23*(0) | 51.8±2.3 | p<0.001 | MNI | SPM99 |
| Li et al. (2006) | In text | 12(8) | 34.56±4.91 | 12(8) | 33.25±5.27 | p<0.001 corrected (ns) | TAL | SPM99 |
| Nardo et al. (2013) | Table 3 | 15(3) | 43.33±8.35 | 17*(6) | 41.59±9.04 | p<0.001 | MNI | SPM5 |
| O'Doherty et al. (2017) | Table 2 | 25(13) | 34±8.4 | 25*(13) | 36.4±8.1 | p<0.001 FWE-corrected | MNI | FSL |
| Qi et al. (2020) | SM | 57(40) | 57.57±5.48 | 163(72) | 58.87±5.53 | p<0.05 FWE-corrected voxel-wise | MNI | SPM12 |
| Rocha-Rego et al. (2012) | Table 2 | 16(9) | 43.4±5.78 | 16*(9) | 44.9±6.6 | p<0.05 FDR-corrected voxel-wise | MNI | SPM5 |
| Sui et al. (2010) | Table 2 | 11(11) | 25.55±4.01 | 12*(8) | 27.5±4 | p<0.05 corrected (ns) | MNI | SPM2 |
| Tan et al. (2013) | Table 2 | 12(0) | 37.6±3.7 | 14*(0) | 40.9±5.2 | p<0.001 | MNI | SPM8 |
| Tavanti et al. (2012) | Table 3 | 25(17) | 38.08±11.01 | 25(17) | 38.16±10.9 | p<0.05 cluster-level corrected (permutation-based) | MNI | FSL 4 |
| Thomaes et al. (2010) | Table 3 | 33(33) | 35.3±9.8 | 30(30) | 35.2±12.3 | p<0.001 | MNI | SPM5 |
| Zhang et al. (2011) | Table 2 | 10(ns) | 40.8±6.83 | 10*(ns) | 34.3±5.37 | p<0.05 corrected (ns) | MNI | SPM2 |
| Zhang et al. (2018a) | Table 2 | 35(17) | 48.22±6.75 | 36*(21) | 50.86±6.62 | p<0.01 FWE-corrected | MNI | SPM8 |

**Table S4:** Records included in the PTSD dataset and their reported medication status.

| PTSD DATASET |  |
| --- | --- |
| Bossini et al. (2017) | medication-free |
| Chao et al. (2012) | antidepressants |
| Cheng et al. (2015) | medication-free |
| Chen et al. (2009) | medication-free |
| Chen et al. (2012) | medication-free |
| Corbo et al. (2005) | medication-free |
| Eckart et al. (2011) | antidepressants |
| Herringa et al. (2012) | medication-free |
| Kasai et al. (2008) | not reported |
| Li et al. (2006) | medication-free |
| Nardo et al. (2013) | medication-free |
| O'Doherty et al. (2017) | antidepressants |
| Qi et al. (2020) | medication-free |
| Rocha-Rego et al. (2012) | antidepressants |
| Sui et al. (2010) | medication-free |
| Tan et al. (2013) | medication-free |
| Tavanti et al. (2012) | medication-free |
| Thomaes et al. (2010) | medication-free |
| Zhang et al. (2011) | medication-free |
| Zhang et al. (2018a) | medication-free |

**Table S5:** Resting-state functional connectivity of the seed at the reported peak coordinates obtained from the ChP meta-analysis.

| CHRONIC PAIN DATASET, seed in [2 -30 40] |  |  |  |
| --- | --- | --- | --- |
| # voxels | Peak Coordinates | Anatomical regions | p-FDR |
| 4398 | [4 -26 38] | Precuneous Cortex; Cingulate Gyrus, posterior division; Hippocampus r; Hippocampus l; Cingulate Gyrus, anterior division; Cuneal Cortex r; Thalamus r; Thalamus l; Cuneal Cortex l | 0.000 |
| 1982 | [8 46 -6] | Cingulate Gyrus, anterior division; Paracingulate Gyrus r; r Frontal Pole r; Subcallosal Cortex; Paracingulate Gyrus l; Superior Frontal Gyrus r; Accumbens l | 0.000 |
| 1393 | [-26 -2 66] | Superior Frontal Gyrus r; Superior Frontal Gyrus l; Juxtapositional Lobule Cortex -formerly Supplementary Motor Cortex- r; Precentral Gyrus l; Juxtapositional Lobule Cortex -formerly Supplementary Motor Cortex- l; Middle Frontal Gyrus l; Precentral Gyrus r | 0.000 |
| 1495 | [50 -62 44] | sLOC r Lateral Occipital Cortex, superior division r; AG r Angular Gyrus r; pSMG r Supramarginal Gyrus, posterior division r | 0.000 |
| 1376 | [-46 -60 48] | Lateral Occipital Cortex, superior division l; Angular Gyrus l; Supramarginal Gyrus, posterior division l; Superior Parietal Lobule l | 0.000 |
| 1168 | [-68 -26 32] | Supramarginal Gyrus, posterior division l; Lateral Occipital Cortex, inferior division l; Supramarginal Gyrus, anterior division l; Middle Temporal Gyrus, temporooccipital part l; Postcentral Gyrus l; Parietal Operculum Cortex l; Angular Gyrus l | 0.000 |
| 1258 | [62 -26 32] | aSMG r Supramarginal Gyrus, anterior division r; SPL r Superior Parietal Lobule r; PostCG r Postcentral Gyrus r; sLOC r Lateral Occipital Cortex, superior division r; pSMG r Supramarginal Gyrus, posterior division r; PO r Parietal Operculum Cortex r | 0.000 |
| 720 | [-42 6 0] | Frontal Orbital Cortex l; Insular Cortex l; Inferior Frontal Gyrus, pars triangularis l; Frontal Operculum Cortex l; Inferior Frontal Gyrus, pars opercularis l; Central Opercular Cortex l | 0.000 |
| 609 | [50 -60 -2] | iLOC r Lateral Occipital Cortex, inferior division r; toITG r Inferior Temporal Gyrus, temporooccipital part r; toMTG r Middle Temporal Gyrus, temporooccipital part r; TOFusC r Temporal Occipital Fusiform Cortex r | 0.000 |
| 617 | [-34 -60 -38] | Cereb2 l Cerebellum Crus2 l; Cereb1 l Cerebellum Crus1 l; Cereb7 l Cerebellum 7b l | 0.000 |
| 477 | [66 -38 -18] | pMTG r Middle Temporal Gyrus, posterior division r toMTG r Middle Temporal Gyrus, temporooccipital part r; pITG r Inferior Temporal Gyrus, posterior division r; toITG r Inferior Temporal Gyrus, temporooccipital part r | 0.000 |
| 458 | [44 16 56] | MidFG r Middle Frontal Gyrus r; SFG r Superior Frontal Gyrus r | 0.000 |
| 308 | [-56 -42 -16] | toITG l Inferior Temporal Gyrus, temporooccipital part l; pMTG l Middle Temporal Gyrus, posterior division l; pITG l Inferior Temporal Gyrus, posterior division l; toMTG l Middle Temporal Gyrus, temporooccipital part l | 0.000 |
| 289 | [-36 16 56] | MidFG l Middle Frontal Gyrus l; SFG l Superior Frontal Gyrus l | 0.000 |
| 313 | [50 -64 -42] | Cerebellum Crus2 r; Cerebellum Crus1 r | 0.000 |
| 189 | [48 2 50] | Precentral Gyrus r | 0.000 |
| 209 | [-54 6 18] | Precentral Gyrus l; Inferior Frontal Gyrus, pars opercularis l | 0.000 |
| 180 | [48 38 32] | Frontal Pole r; Middle Frontal Gyrus r | 0.000 |
| 100 | [54 36 4] | Frontal Pole r; Inferior Frontal Gyrus, pars triangularis r | 0.000 |
| 161 | [-14 -76 -50] | Cerebellum 8 l; Cerebellum 7b l; Cerebellum Crus2 l | 0.000 |
| 129 | [40 0 12] | Central Opercular Cortex r; IC r Insular Cortex r | 0.000 |
| 147 | [-40 0 -38] | Inferior Temporal Gyrus, anterior division l; Temporal Fusiform Cortex, anterior division l | 0.000 |
| 137 | [-40 -50 -18] | Temporal Fusiform Cortex, posterior division l; Temporal Occipital Fusiform Cortex l | 0.000 |
| 142 | [-18 -56 72] | Superior Parietal Lobule l | 0.000 |
| 130 | [42 54 12] | Frontal Pole r | 0.000 |
| 126 | [-36 54 -2] | Frontal Pole l | 0.000 |
| 71 | [36 16 -6] | Insular Cortex r | 0.000 |
| 35 | [2 -18 -18] | Brain-Stem | 0.001 |
| 58 | [24 48 -12] | Frontal Pole r | 0.002 |
| 47 | [-44 4 -28] | Temporal Pole l | 0.002 |
| 48 | [22 -74 -36] | Cerebellum Crus2 r | 0.003 |
| 48 | [48 10 -20] | Temporal Pole r | 0.004 |
| 31 | [-8 -22 16] | Thalamus l | 0.006 |
| 44 | [22 -98 -6] | Occipital Pole r | 0.006 |
| 38 | [-26 34 -20] | Frontal Orbital Cortex l | 0.006 |
| 29 | [14 -26 16] | Thalamus r | 0.018 |
| 28 | [18 30 -24] | Frontal Orbital Cortex r | 0.021 |
| 26 | [-22 46 -10] | Frontal Pole l | 0.027 |
| 27 | [24 -68 -56] | Cerebellum 8 r | 0.030 |

**Table S6:** Resting-state functional connectivity of the seed at the reported peak coordinates obtained from the ChP meta-analysis.

| CHRONIC PAIN DATASET, seed in [-6 -48 -60] |  |  |  |
| --- | --- | --- | --- |
| # voxels | Peak Coordinates | Anatomical regions | p-FDR |
| 7565 | [-6 50 -2] | Precuneus Cortex; Cingulate Gyrus, posterior division; Subcallosal Cortex; Cingulate Gyrus, anterior division; Paracingulate Gyrus l; Frontal Pole l; Frontal Medial Cortex; Paracingulate Gyrus r; Hippocampus l; Hippocampus r; Frontal Pole r; Thalamus l; Parahippocampal Gyrus, posterior division l; Thalamus r; r Parahippocampal Gyrus, posterior division r; Accumbens l; Accumbens r; Lingual Gyrus l; Parahippocampal Gyrus, anterior division l | 0.000 |
| 666 | [46 28 10] | Inferior Frontal Gyrus, pars opercularis r; Inferior Frontal Gyrus, pars triangularis r; Frontal Pole r; Frontal Operculum Cortex r; Middle Frontal Gyrus r | 0.000 |
| 687 | [40 54 -4] | Frontal Pole r | 0.000 |
| 635 | [54 -38 56] | Supramarginal Gyrus, posterior division r; Angular Gyrus r; Superior Parietal Lobule r; Supramarginal Gyrus, anterior division r | 0.000 |
| 639 | [-8 -84 -22] | Cerebellum Crus1 l; Cerebellum Crus2 l; Cerebellum 6 l | 0.000 |
| 419 | [-58 -10 -14] | Middle Temporal Gyrus, posterior division l; Middle Temporal Gyrus, anterior division l; Superior Temporal Gyrus, anterior division l | 0.000 |
| 423 | [-52 -72 30] | Lateral Occipital Cortex, superior division l; Angular Gyrus l | 0.000 |
| 341 | [-18 42 46] | Superior Frontal Gyrus l; Frontal Pole l | 0.000 |
| 232 | [6 30 48] | Superior Frontal Gyrus r; r Paracingulate Gyrus r | 0.000 |
| 267 | [-44 48 4] | Frontal Pole l | 0.000 |
| 132 | [-30 -78 -44] | Cerebellum Crus2 l; Cerebellum 7b l | 0.000 |
| 153 | [48 26 46] | Middle Frontal Gyrus r | 0.000 |
| 132 | [32 56 -18] | Frontal Pole r | 0.000 |
| 148 | [6 -48 -42] | Cerebellum 9 r | 0.000 |
| 103 | [72 -40 0] | Middle Temporal Gyrus, temporooccipital part r | 0.000 |
| 113 | [26 -68 -28] | Cerebellum 6 r; Cerebellum Crus1 r | 0.000 |
| 85 | [62 -8 -12] | Middle Temporal Gyrus, anterior division r; pMiddle Temporal Gyrus, posterior division r | 0.000 |
| 50 | [-62 -26 28] | Supramarginal Gyrus, anterior division l | 0.000 |
| 48 | [-52 12 -4] | Inferior Frontal Gyrus, pars opercularis l | 0.000 |
| 56 | [-38 -48 50] | Superior Parietal Lobule l | 0.001 |
| 44 | [-46 10 16] | Inferior Frontal Gyrus, pars opercularis l | 0.003 |
| 26 | [42 12 60] | Middle Frontal Gyrus r | 0.013 |

**Table S7:** Resting-state functional connectivity of the seed at the reported peak coordinates obtained from the ChP meta-analysis.

| CHRONIC PAIN DATASET, seed in [32 16 -12] |  |  |  |
| --- | --- | --- | --- |
| # voxels | Peak Coordinates | Anatomical regions | p-FDR |
| 4063 | [4 36 10] | Cingulate Gyrus, anterior division; Paracingulate Gyrus r; Superior Frontal Gyrus r; Paracingulate Gyrus l; Subcallosal Cortex; Frontal Pole r; Superior Frontal Gyrus l | 0.000 |
| 1666 | [32 18 -14] | Frontal Orbital Cortex r; Insular Cortex r; Inferior Frontal Gyrus, pars triangularis r; Frontal Operculum Cortex r; Inferior Frontal Gyrus, pars opercularis r; Putamen r; Caudate r | 0.000 |
| 1170 | [64 0 34] | Postcentral Gyrus r; Precentral Gyrus r; Superior Parietal Lobule r; Central Opercular Cortex r; Supramarginal Gyrus, anterior division r | 0.000 |
| 659 | [-26 16 -16] | Frontal Orbital Cortex l; Insular Cortex l; Putamen l; Accumbens l | 0.000 |
| 640 | [56 -48 46] | Angular Gyrus r; Supramarginal Gyrus, posterior division r | 0.000 |
| 458 | [4 -20 36] | Cingulate Gyrus, posterior division; Cingulate Gyrus, anterior division | 0.000 |
| 483 | [-54 -10 40] | Postcentral Gyrus l; Precentral Gyrus l; Central Opercular Cortex l | 0.000 |
| 491 | [26 62 24] | Frontal Pole r | 0.000 |
| 331 | [56 -34 -6] | Middle Temporal Gyrus, posterior division r | 0.000 |
| 377 | [44 10 44] | Middle Frontal Gyrus r | 0.000 |
| 279 | [6 -10 -2] | Thalamus r; Thalamus l | 0.000 |
| 345 | [12 -66 34] | Precuneus Cortex; Cingulate Gyrus, posterior division | 0.000 |
| 271 | [26 -8 72] | Precentral Gyrus r; Superior Frontal Gyrus r | 0.000 |
| 297 | [28 58 -10] | Frontal Pole r | 0.000 |
| 261 | [-58 -52 36] | Supramarginal Gyrus, posterior division l; Angular Gyrus l | 0.000 |
| 195 | [-24 -50 68] | Superior Parietal Lobule l; Postcentral Gyrus l | 0.000 |
| 175 | [50 8 -36] | Temporal Pole r; Middle Temporal Gyrus, anterior division r | 0.000 |
| 165 | [-30 -6 70] | Superior Frontal Gyrus l; Precentral Gyrus l; SJuxtapositional Lobule Cortex - formerly Supplementary Motor Cortex- l | 0.000 |
| 129 | [-60 -44 -6] | Middle Temporal Gyrus, temporooccipital part l | 0.000 |
| 183 | [-48 -70 18] | Lateral Occipital Cortex, superior division l | 0.000 |
| 184 | [-24 -82 -32] | Cerebellum Crus1 l; Cereb2 l Cerebellum Crus2 l | 0.000 |
| 102 | [-38 42 -4] | Frontal Pole l | 0.000 |
| 105 | [-44 30 16] | Inferior Frontal Gyrus, pars triangularis l; Frontal Pole l | 0.000 |
| 100 | [28 -24 -12] | Hippocampus r | 0.000 |
| 71 | [-8 -68 44] | Precuneus Precuneus Cortex | 0.000 |
| 69 | [-10 22 68] | Superior Frontal Gyrus l | 0.000 |
| 65 | [-24 56 28] | Frontal Pole l | 0.001 |
| 66 | [-64 -28 -8] | Middle Temporal Gyrus, posterior division l | 0.001 |
| 33 | [-42 -48 -24] | Inferior Temporal Gyrus, temporooccipital part l | 0.001 |
| 58 | [-48 -58 -44] | Cerebellum Crus2 l | 0.001 |
| 32 | [56 -28 20] | Supramarginal Gyrus, anterior division r | 0.002 |
| 26 | [-26 22 48] | Middle Frontal Gyrus l | 0.004 |
| 38 | [-54 -66 -36] | l Cerebellum Crus1 l | 0.009 |
| 30 | [2 -18 -18] | Brain-Stem | 0.012 |
| 33 | [-28 -76 52] | Lateral Occipital Cortex, superior division l | 0.017 |
| 26 | [-36 -42 56] | Superior Parietal Lobule l | 0.029 |
| 25 | [-54 -36 24] | Parietal Operculum Cortex l | 0.042 |

**Table S8:** Resting-state functional connectivity of the seed at the reported peak coordinates obtained from the ChP meta-analysis.

| CHRONIC PAIN DATASET, seed in [-50 -20 12] |  |  |  |
| --- | --- | --- | --- |
| # voxels | Peak Coordinates | Anatomical regions | p-FDR |
| 26102 | [-46 -20 20] | Postcentral Gyrus l; Precentral Gyrus l; Postcentral Gyrus r; Precentral Gyrus r; Central Opercular Cortex l; Insular Cortex r; Insular Cortex l; Central Opercular Cortex r; Cingulate Gyrus, anterior division; Juxtapositional Lobule Cortex -formerly Supplementary Motor Cortex- r; Planum Temporale l; Parietal Operculum Cortex l; Juxtapositional Lobule Cortex -formerly Supplementary Motor Cortex- l; Parietal Operculum Cortex r; Superior Parietal Lobule l; Superior Parietal Lobule r; Planum Temporale r; Supramarginal Gyrus, anterior division r; Superior Frontal Gyrus r; PP r Planum Polare r; Planum Polare l; Heschl's Gyrus l; Heschl's Gyrus r; Superior Frontal Gyrus l; Superior Temporal Gyrus, posterior division r; Supramarginal Gyrus, anterior division l; Superior Temporal Gyrus, posterior division l; Temporal Pole l; Cingulate Gyrus, posterior division; Temporal Pole r; Precuneous Precuneous Cortex; Superior Temporal Gyrus, anterior division l; Paracingulate Gyrus r; Superior Temporal Gyrus, anterior division r; Putamen l; Paracingulate Gyrus l; Frontal Operculum Cortex r; Frontal Operculum Cortex l; Inferior Frontal Gyrus, pars opercularis l; Supramarginal Gyrus, posterior division r; Putamen r | 0.000 |
| 2160 | [12 -60 -20] | Cerebelum 6 r; Cerebelum 6 l; Lingual Gyrus r; Intracalcarine Cortex l; Cerebelum 4 5 r; Lingual Gyrus l; Cerebelum 4 5 l; Vermis 6; Vermis 4 5; Intracalcarine Cortex r; Cuneal l Cuneal Cortex l; Precuneous Precuneous Cortex; Hippocampus r; Supracalcarine Cortex r; Cuneal r Cuneal Cortex r | 0.000 |
| 882 | [28 -52 -48] | Cerebelum 8 r; Cerebelum 8 l; Vermis 8; Cerebelum 9 l | 0.000 |
| 572 | [-18 -68 -32] | Cerebelum Crus1 l; Cerebelum Crus2 l; Cerebelum 7b l; Cerebelum 6 l | 0.000 |
| 554 | [-26 -76 42] | Lateral Occipital Cortex, superior division l | 0.000 |
| 490 | [-50 -62 -14] | Inferior Temporal Gyrus, temporooccipital part l; Middle Temporal Gyrus, temporooccipital part l; Lateral Occipital Cortex, inferior division l | 0.000 |
| 514 | [28 -64 -30] | Cerebelum Crus1 r; Cerebelum 6 r; Cerebelum Crus2 r | 0.000 |
| 329 | [14 -18 0] | Thalamus r | 0.000 |
| 351 | [-16 -24 8] | Thalamus l | 0.000 |
| 343 | [46 -78 24] | Lateral Occipital Cortex, superior division r | 0.000 |
| 165 | [-44 -48 52] | Supramarginal Gyrus, posterior division l | 0.000 |
| 176 | [-2 -68 60] | Precuneous Precuneous Cortex; Lateral Occipital Cortex, superior division l | 0.000 |
| 101 | [56 26 -2] | Frontal Pole r; Inferior Frontal Gyrus, pars triangularis r | 0.000 |
| 121 | [-26 16 64] | Superior Frontal Gyrus l | 0.000 |
| 84 | [14 -84 32] | Cuneal Cortex r | 0.000 |
| 73 | [12 18 70] | Superior Frontal Gyrus r | 0.000 |
| 28 | [2 -50 -54] | Cerebelum 9 r | 0.000 |
| 47 | [34 -74 -56] | Cerebelum 7b r | 0.000 |
| 70 | [30 62 16] | Frontal Pole r | 0.001 |
| 60 | [52 -54 24] | Angular Gyrus r | 0.001 |
| 47 | [28 -74 52] | Lateral Occipital Cortex, superior division r | 0.005 |
| 43 | [6 -22 -42] | Brain-Stem | 0.006 |
| 39 | [-28 46 -18] | Frontal Pole l | 0.008 |
| 33 | [-48 -54 -48] | Cerebelum Crus2 l | 0.008 |
| 28 | [-32 24 8] | Frontal Operculum Cortex l | 0.009 |
| 26 | [-22 -34 -40] | Cerebelum 10 l | 0.012 |
| 25 | [46 30 -20] | Frontal Orbital Cortex r | 0.015 |
| 34 | [-8 -30 -40] | Brain-Stem | 0.015 |
| 25 | [-4 -82 -22] | Cerebelum Crus2 l | 0.023 |
| 25 | [-30 58 14] | Frontal Pole l | 0.041 |

**Table S9:**
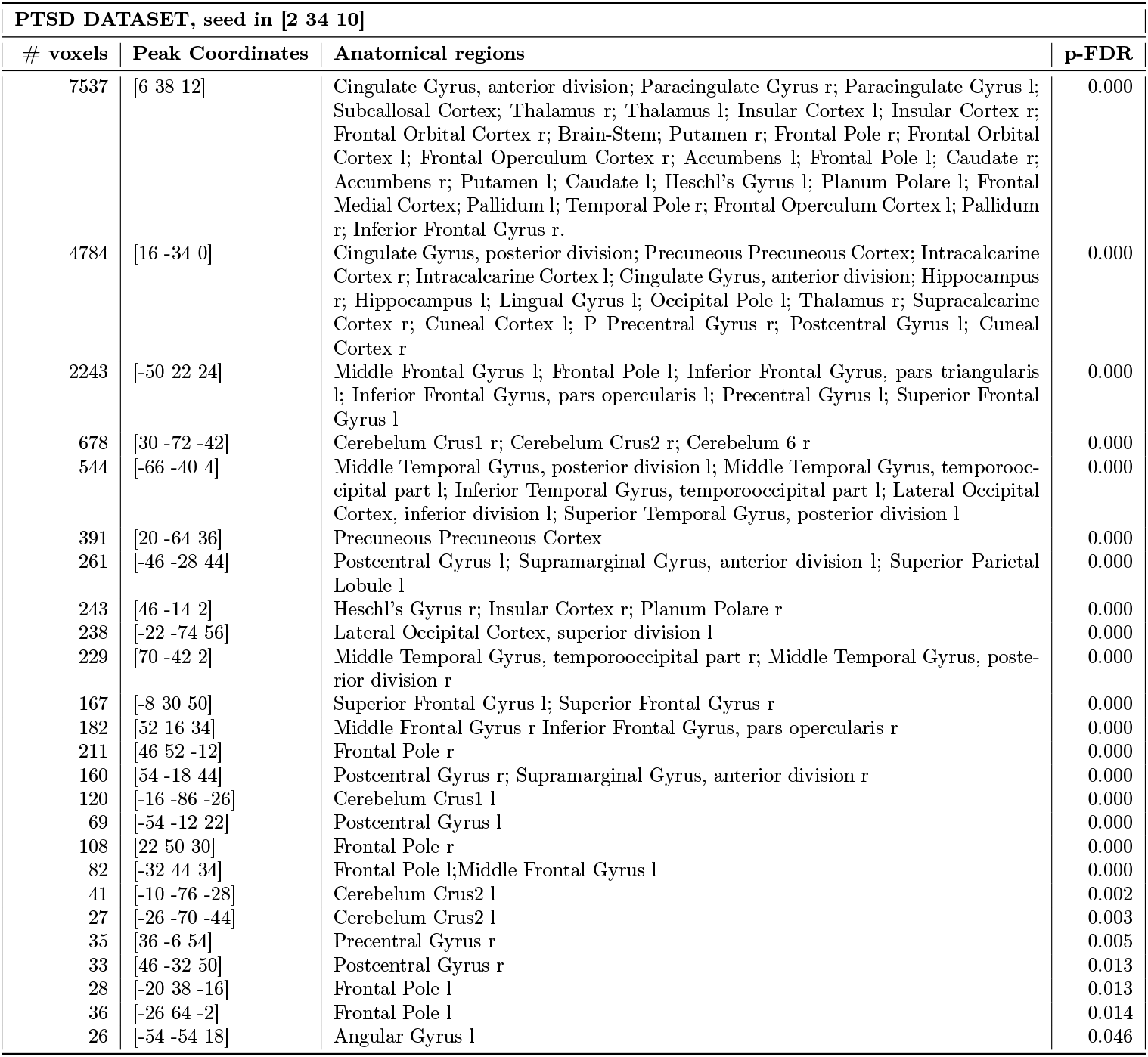
Resting-state functional connectivity of the seed at the reported peak coordinates obtained from the PTSD meta-analysis.

| PTSD DATASET, seed in [2 34 10] |  |  |  |
| --- | --- | --- | --- |
| # voxels | Peak Coordinates | Anatomical regions | p-FDR |
| 7537 | [6 38 12] | Cingulate Gyrus, anterior division; Paracingulate Gyrus r; Paracingulate Gyrus l; Subcallosal Cortex; Thalamus r; Thalamus l; Insular Cortex l; Insular Cortex r; Frontal Orbital Cortex r; Brain-Stem; Putamen r; Frontal Pole r; Frontal Orbital Cortex l; Frontal Operculum Cortex r; Accumbens l; Frontal Pole l; Caudate r; Accumbens r; Putamen l; Caudate l; Heschl's Gyrus l; Planum Polare l; Frontal Medial Cortex; Pallidum l; Temporal Pole r; Frontal Operculum Cortex l; Pallidum r; Inferior Frontal Gyrus r. | 0.000 |
| 4784 | [16 -34 0] | Cingulate Gyrus, posterior division; Precuneous Precuneous Cortex; Intracalcarine Cortex r; Intracalcarine Cortex l; Cingulate Gyrus, anterior division; Hippocampus r; Hippocampus l; Lingual Gyrus l; Occipital Pole l; Thalamus r; Supracalcarine Cortex r; Cuneal Cortex l; P Precentral Gyrus r; Postcentral Gyrus l; Cuneal Cortex r | 0.000 |
| 2243 | [-50 22 24] | Middle Frontal Gyrus l; Frontal Pole l; Inferior Frontal Gyrus, pars triangularis l; Inferior Frontal Gyrus, pars opercularis l; Precentral Gyrus l; Superior Frontal Gyrus l | 0.000 |
| 678 | [30 -72 -42] | Cerebellum Crus1 r; Cerebellum Crus2 r; Cerebellum 6 r | 0.000 |
| 544 | [-66 -40 4] | Middle Temporal Gyrus, posterior division l; Middle Temporal Gyrus, temporooccipital part l; Inferior Temporal Gyrus, temporooccipital part l; Lateral Occipital Cortex, inferior division l; Superior Temporal Gyrus, posterior division l | 0.000 |
| 391 | [20 -64 36] | Precuneous Precuneous Cortex | 0.000 |
| 261 | [-46 -28 44] | Postcentral Gyrus l; Supramarginal Gyrus, anterior division l; Superior Parietal Lobule l | 0.000 |
| 243 | [46 -14 2] | Heschl's Gyrus r; Insular Cortex r; Planum Polare r | 0.000 |
| 238 | [-22 -74 56] | Lateral Occipital Cortex, superior division l | 0.000 |
| 229 | [70 -42 2] | Middle Temporal Gyrus, temporooccipital part r; Middle Temporal Gyrus, posterior division r | 0.000 |
| 167 | [-8 30 50] | Superior Frontal Gyrus l; Superior Frontal Gyrus r | 0.000 |
| 182 | [52 16 34] | Middle Frontal Gyrus r Inferior Frontal Gyrus, pars opercularis r | 0.000 |
| 211 | [46 52 -12] | Frontal Pole r | 0.000 |
| 160 | [54 -18 44] | Postcentral Gyrus r; Supramarginal Gyrus, anterior division r | 0.000 |
| 120 | [-16 -86 -26] | Cerebellum Crus1 l | 0.000 |
| 69 | [-54 -12 22] | Postcentral Gyrus l | 0.000 |
| 108 | [22 50 30] | Frontal Pole r | 0.000 |
| 82 | [-32 44 34] | Frontal Pole l; Middle Frontal Gyrus l | 0.000 |
| 41 | [-10 -76 -28] | Cerebellum Crus2 l | 0.002 |
| 27 | [-26 -70 -44] | Cerebellum Crus2 l | 0.003 |
| 35 | [36 -6 54] | Precentral Gyrus r | 0.005 |
| 33 | [46 -32 50] | Postcentral Gyrus r | 0.013 |
| 28 | [-20 38 -16] | Frontal Pole l | 0.013 |
| 36 | [-26 64 -2] | Frontal Pole l | 0.014 |
| 26 | [-54 -54 18] | Angular Gyrus l | 0.046 |

**Table S10:** Resting-state functional connectivity of the seed at the reported peak coordinates obtained from the conjunction analysis.

| Conjunction, seed in [2 -30 40] |  |  |  |
| --- | --- | --- | --- |
| # voxels | Peak Coordinates | Anatomical regions | p-FDR |
| 4015 | [0 46 -10] | Subcallosal Cortex; Frontal Medial Cortex; Cingulate Gyrus, anterior division; Paracingulate Gyrus l; Frontal Pole l; PaCiG r Paracingulate Gyrus r; Frontal Pole r; Accumbens l; Accumbens r | 0.000 |
| 3811 | [-2 -56 24] | Precuneous Cortex; Cingulate Gyrus, posterior division; Hippocampus l; Hippocampus r Parahippocampal Gyrus, posterior division l; Parahippocampal Gyrus, posterior division r; Thalamus l; Amygdala r; Lingual Gyrus l; Vermis 4 5; Thalamus r; Amygdala l | 0.000 |
| 884 | [56 24 10] | Inferior Frontal Gyrus, pars opercularis r; Inferior Frontal Gyrus, pars triangularis r; Frontal Pole r; Middle Frontal Gyrus r | 0.000 |
| 920 | [56 -40 46] | Supramarginal Gyrus, posterior division r; Angular Gyrus r; Supramarginal Gyrus, anterior division r; Superior Parietal Lobule r | 0.000 |
| 887 | [42 44 -2] | Frontal Pole r; Frontal Orbital Cortex r | 0.000 |
| 808 | [-40 -48 48] | Supramarginal Gyrus, posterior division l; Supramarginal Gyrus, anterior division l; Superior Parietal Lobule l; Angular Gyrus l | 0.000 |
| 606 | [0 32 48] | Superior Frontal Gyrus r; Superior Frontal Gyrus l; Paracingulate Gyrus r; Paracingulate Gyrus l; Juxtapositional Lobule Cortex -formerly Supplementary Motor Cortex- l | 0.000 |
| 660 | [-42 48 4] | Frontal Pole l; Middle Frontal Gyrus l | 0.000 |
| 481 | [-58 -14 -14] | Middle Temporal Gyrus, posterior division l Middle Temporal Gyrus, anterior division l; Superior Temporal Gyrus, anterior division l | 0.000 |
| 597 | [-18 -74 -20] | Cerebellum Crus1 l; Cerebellum Crus2 l; Cerebellum 6 l | 0.000 |
| 402 | [-40 -64 30] | Lateral Occipital Cortex, superior division l | 0.000 |
| 340 | [46 26 46] | Middle Frontal Gyrus r | 0.000 |
| 227 | [-56 16 -2] | Inferior Frontal Gyrus, pars opercularis l; Insular Cortex l | 0.000 |
| 263 | [-22 38 46] | Superior Frontal Gyrus l; Frontal Pole l | 0.000 |
| 278 | [26 -68 -28] | Cerebellum Crus1 r; Cerebellum 6 r; Cereb2 r Cerebellum Crus2 r | 0.000 |
| 253 | [04 -46 -46] | Cerebellum 9 r; Cerebellum 9 l | 0.000 |
| 212 | [64 -4 -14] | Middle Temporal Gyrus, anterior division r; Middle Temporal Gyrus, posterior division r | 0.000 |
| 173 | [-40 0 40] | Middle Frontal Gyrus l; Precentral Gyrus l | 0.000 |
| 86 | [-26 -72 -52] | Cerebellum Crus2 l; Cerebellum 8 l | 0.000 |
| 120 | [24 -74 -44] | Cerebellum 8 r; Cerebellum 7b r | 0.000 |
| 123 | [2 -14 6] | Thalamus l; Thalamus r | 0.000 |
| 73 | [-12 14 72] | Superior Frontal Gyrus l | 0.000 |
| 31 | [36 34 -8] | Frontal Pole r | 0.003 |
| 35 | [46 -76 -40] | Cerebellum Crus2 r | 0.009 |
| 36 | [-32 30 -16] | Frontal Orbital Cortex l | 0.011 |
| 31 | [-24 -62 44] | Lateral Occipital Cortex, superior division l | 0.012 |
| 25 | [-20 42 -18] | Frontal Pole l | 0.023 |

